# Perceived Clinical Readiness of Senior Medical Students as Outcomes of Online Clerkship in the Philippines: New Normal in Medical Education

**DOI:** 10.1101/2023.05.11.23289727

**Authors:** Adam James A. Abear, Marie Bernice P. Benitez, Krissha Marie S. Cabrillos, Adrian V. Casaña, Ma. Theresa Mae P. Doctora, Joan Marie D. Ellema, Bianca Louise U. Fuentes, Justin Riley Y. Lam, John Emmanuel C. Mendoza, Shairah P. Tan, Shannon D. Tan

## Abstract

**TITLE:** Perceived Clinical Readiness of Senior Medical Students as Outcomes 0f Online Clerkship in the Philippines: New Normal in Medical Education

**INTRODUCTION:** COVID-19 formed new challenges to the medical institutions; it resulted in the transition from the usual face-to-face classes and direct clerkship training within the hospital to a new remote learning with online lectures and virtual clinical experience. Given the new online interactive setting, problems were raised given the limited patient care and interaction as well as restricted bedside teaching opportunities and its impact on how medical students can acquire and hone their clinical skills.

**OBJECTIVE:** To determine the perceived clinical readiness of the medical clerks in the new normal setting in the Philippines.

**METHODOLOGY:** Convenience sampling was used to gather respondents who were asked to answer an online survey questionnaire. The questions pertained to: academic training profile, clinical skills, patient management, communication, understanding clinical guidelines, and personal development. After analyzing the data, the scales of readiness from these subjects were gathered.

**RESULTS:** The medical clerks in the Philippines perceived that they were ready with regards to understanding clinical guidelines, communication, personal development, and patient management. They were moderately ready in the different clinical skills in the departments of Family and Community Medicine, Internal Medicine, Pediatrics, Surgery and Obstetrics and Gynecology with some specific skills in Surgery and Obstetrics and Gynecology being perceived as less ready than the rest.

**CONCLUSION:** The impact of the pandemic has disrupted the student’s confidence and readiness. This shows that online clerkship in this time of pandemic may have provided learning to a certain degree but it is not enough to replace what face-to-face training could offer.

## INTRODUCTION

### Background of the study

As COVID-19 posed a deadly threat, medical institutions are now faced with new challenges in delivering and continuing the medical education process safely while maintaining its standards.^1^ This has resulted in a transition from the usual face-to-face classes and direct clerkship training within a hospital to a new platform of remote learning with online lectures and conferences, and virtual clinical experience. Medical schools have temporarily suspended their clerkship rotation and training in a hospital setting to limit the spread of COVID-19 infection by minimizing personal interaction and in turn decreasing the risk of exposure of medical students.

The use of online interactive discussions and virtual clinical settings may allow the students to perfect the educational theory and exhibit the competencies necessary for progression and continue their training.^2^ Virtual web-based platforms such as Zoom and Google Meet have been used for the delivery of webinars, case discussions, reading assignments, and small group discussions (SGDs, peer mentoring). Online learning has also been shown to be as effective as conventional teaching because it promotes self-directed learning,^3^ develops interactive virtual clinical teaching, and encourages student engagement and student feedback, with added benefits of easy accessibility of learning materials.^4^

However limited patient care and interaction can restrict bedside teaching opportunities and can impact how medical students acquire and hone their clinical skills, and develop collaborative experiences. Also, such measures being adapted cannot wholly replace the physician-to-patient interaction which is integral to medical training.^1^ Moreover, there has been a concern that medical students may miss out on the experiences of clinical rotations, such as exposure to an authentic patient environment. This may be a challenge since the students are encouraged to be innovative and adaptive in developing and demonstrating their skills and knowledge. However, this situation may also provide a way for the students and medical educators to analyze the outcome of the recent changes in the medical set-up and provide new principles and practices that may contribute to the advancement of medical education.^5^ In this study, the researchers aimed to provide an overview of medical students’ clinical readiness during this time of the pandemic where digital medical education is being instituted, and alternative clinical clerkship training programs and new modes of student-patient interaction are practiced.

### Significance of the study

Meeting the competencies and completing the requirements are needed to progress in the clinical clerkship training and to become a well-rounded physician in the future. With the sudden transition to remote learning brought by the pandemic, various aspects of the medical education process have been affected, with much of the impact seen on the medical students who had to ensure they acquire the necessary clinical skills and experience while they continue to adapt to the new platform. The findings of this study will be of considerable importance to the following population:

#### Medical students

This study will allow medical students to evaluate what clinical skills they have acquired so far in the practice of medicine and to assess their level of mastery and confidence when asked to perform the following procedures during return demonstration and in the actual hospital setting. From there, students can have an insight into what areas need improvement and how to overcome the challenges of online learning.

#### Medical schools

Determining the impact of the alternative clerkship training programs on the clinical readiness of the medical students can guide medical schools on what to modify on their current strategies for curriculum delivery and how to formulate new ones to improve clinical skills teaching and training experience through the adapted online platform.

#### The community

Conducting this study will also make the community aware that containment of the virus and rapid decline of cases not only aids the healthcare workers but also encompasses the rehabilitation of different sectors, most especially, the education department. This will allow the continuation of physician-to-patient interaction and honing of fundamental hands-on clinical skills.

### Research Question

How does online clinical clerkship training programs affect the preparedness of the fourth year medical students in the Philippines to practice medicine?

### General Objectives

The study aims to determine the level of clinical readiness of the fourth year medical students in the Philippines with regards to the performance of clinical skills under the new online learning platform in the practice of medicine.

### Specific Objectives

Specifically, this study seeks to answer the following questions:

1. Academic training profile of the students
  1.1 Performance last academic year
  1.2 Suspension of face-to-face clinical rotations
  1.3 Length of time with face-to-face experience
  1.4 Types of online activities
  1.5 Methods of evaluation for skills
  1.6 Learning management system
2. What is the extent of readiness as an impact of online clinical training on the developmental areas of:
  2.1 Clinical Skills
    2.1-1 General Clinical Skills
    2.1-2 Department of Obstetrics and Gynecology
    2.1-3 Department of Surgery
    2.1-4 Department of Internal Medicine
    2.1-5 Department of Pediatrics
    2.1-6 Department of Family and Community Medicine
  2.2 Patient Management
  2.3 Communication and Team Working
  2.5 Understanding Clinical Guidelines and protocols
  2.6 Personal development and wellbeing
3. Is there a significant difference in the readiness level of the respondents with respect to the different mandated preparatory skills?
4. Based on the results, what strategies can be forwarded to improve the online delivery of clinical training?

## RESEARCH METHODOLOGY

### Study Design

This study employed a descriptive research design using the cross-sectional survey method. The researchers measured the clinical preparedness of fourth year medical students in the Philippines, hereinafter referred to as respondents, based on their online clinical learning for this academic year 2020-2021. An online questionnaire was used for the survey.

### Study Setting

This study targeted respondents from the *44* medical schools in the Philippines. The medical schools are found all over the country in different regions namely: Region 1, Region 2, Region 3, Region 4A, MIMAROPA, Region 5, Region 6, Region 7, Region 8, Region 9, Region 10, Region 11, and NCR.

### Study Population

The population of interest was composed of the fourth year medical students, approximately 5,500, in the Philippines with an estimated cohort of 44. Respondents were enrolled in their respective medical schools as fourth year medical students for the academic year 2020-2021.

### Sampling design

This study employed a multi-stage sampling. The first stage was stratified random sampling where the researchers took the medical schools as strata. The second stage was to do convenient sampling due to the nature of availability, commitment, and consent of the students from each school with a targeted minimum of 30 students from each school based on the central limit theorem.

### Inclusion Criteria

The participants included those who were enrolled as regular fourth year medical students or those who were conditionally enrolled (with pending requirements for submission) as long as they received the same training in this case little to no face-to-face classes as the other regular medical students.

### Exclusion Criteria

Participants who were currently on a leave of absence (LOA), those who were irregular students, or those who changed medical schools during their training were excluded from the analysis.

### Data Collection Process

The researchers tapped a student network organization based in the Philippines. From there, the online questionnaire was sent through the “Google Forms” link along with a copy of “Informed Consent,” which was distributed to all medical schools in the Philippines via an organization’s representative, in coordination with the researchers. The said organization is a non-stock, non-profit, and voluntary, that is incorporated under the Philippine laws. With their mission to promote quality and medical education in the country, the researchers chose them because the data that can be collected may serve as a primary source of information about the clinical preparedness of senior clerkship. The researchers then set a time frame for the collection of data and strictly adhered to the schedule. Constant follow-up was done. Responses to the forms were automatically saved in a drive where only the researchers can gain access. Once all the data were collected and collated, the analysis followed in two weeks.

### Data Collection Tool

The researchers crafted a simple survey/questionnaire *(See Appendix A)* focusing on the clinical preparedness of the respondents, based on the competency set nationally by the Commission on Higher Education (CHED). The questionnaire aimed to explore the readiness of interns as they transitioned themselves into graduates in the Doctor of Medicine (M.D.) program, how well they were prepared, and how confident they were in dealing with actual patients in the clinical setting, in the effect of this global pandemic brought about by COVID-19. The *n*-item questionnaire was designed to combine multiple choice, Likert scale questions, and open-ended questions.

### Validity and Reliability

The crafted questionnaire was evaluated and scrutinized by different panels, composed of medical students, undergraduate and postgraduate medical educators, including the Research Adviser and Research Director of the researchers’ academic institution. For the face validity, the research questionnaire satisfactorily met the researchers’ objective in exploring senior clerk’s readiness and preparedness for the clinical rotation, as approved by the panel’s consensus. To ensure content validity, questions were based accordingly on the minimum competencies or program outcomes in the four major areas of clinical clerkship mentioned in the Joint Memorandum Order No. 2016-18. Researchers also did cross-reference with areas explored by other authors in the published literature. Construct validity was not a problem as self-perceptions of the respondents were gathered instead of abstract concepts. The Cronbach Alpha’s Test was used to measure the internal consistency and reliability of the questionnaire, the instrument obtained an excellent internal consistency with a Cronbach alpha value of 0.98 *(See Appendix B)*.

### Statistical Treatment

For the categorical data especially on the profiling, frequency and simple percentage were used. In terms of analyzing the perceived readiness, a weighted mean was computed. Likewise, the Analysis of Variance (One Factorial Design) was employed to determine the significance of the differences of readiness levels across the developmental areas. A Tukey’s post hoc test was applied when significance was detected at 0.05 level of significance.

### Ethical Consideration

The research was subjected to examination and approval by the Institutional Review Board (IRB) of Cebu Institute of Medicine (CIM). A detailed description of the study including the rationale, objectives, and procedures was provided to the student network organization. Any form of risk or harm while answering the survey questionnaire was reported to the student network organization, researchers, and the IRB. The researchers obtained informed consent from the participants before the survey began.

### Informed Consent

This study obtained informed consent *(See Appendix C)* from our respondents that followed the WHO-ERB standards. All the following elements were detailed in the informed consent: the purpose of the study, type of research intervention, participant selection, voluntary participation, procedures, duration, risks, benefits, and confidentiality. As previously mentioned, participation was completely voluntary and the respondent had total freedom to decide whether to finish the questionnaire or not. Strictly, all identifying and personal information remained confidential.

### Confidentiality and Security of Information

The questionnaire was submitted to the IRB of CIM. Before answering the questionnaire, the respondents were made to read and respond to the informed consent that was presented on the first page of the online questionnaire. Participation of the respondents was voluntary. Respondents were assured that the information gathered from the survey was used for research purposes only and that their identities remained anonymous. Information obtained in this study included the personal information of the respondents and was kept confidential. The following measures were instituted:

1. Completed questionnaire did not contain identifying information. They were marked with a serial code and a separate identifier sheet was kept with the corresponding serial code matching the one on the completed questionnaire. The identifier sheet was kept and made known only to the primary investigator. Identifiable data were encrypted.
2. The files were only stored and accessible with the account of the principal investigator
3. All study data were disposed of promptly after the study duration.

### Non-Maleficence

Each participant was informed of the objectives and design of the study in the informed consent forms.

### Beneficence

The results of the study will be used to contribute to novel and existing knowledge in the use of online learning for the senior clerkship or fourth year curriculum.

### Benefits

Since this study did not require compulsory participation of the respondents, they had the right to withdraw from participating in this research study if they wished to do so. They also had the right to ask about the analysis of data gathered from the data collection.

### Protection

Any information that could incriminate the respondents nor any personal data provided was not released without the patient’s consent. The data collectors were third year medical students who were rigorously prepared to handle and administer the questionnaire and address any concerns that would possibly harm the respondents.

## RESULTS

The responses of the senior year medical students in different medical schools in the Philippines were tabulated, analyzed, and interpreted. The first section is on the personal and online learning profile of the students while the second section is on the extent of perceived readiness as an impact of online clinical training on the respective developmental areas, and the third section is about the hypothesis testing for the significance of the mean differences in their perceptions with respect to the different mandated preparatory skills.

### Section 1: Personal and online learning profile of the respondents

Table 1 shows the personal and learning profile of the respondents where it can be verified that most senior medical students who participated in the survey were females (62.43%), with a mean age of 25.85 years old. Almost all respondents (98.80%) also revealed that there was a suspension of face-to-face clinical rotations due to the COVID-19 pandemic. A majority did not also plan for a leave of absence (58.56%) where 62.34% claimed to have had face-to-face duty during the worldwide health crisis. For those who had experiences of face-to-face duties, 61% of them claimed that they were on duty for only less than a month.

**Table 1.**
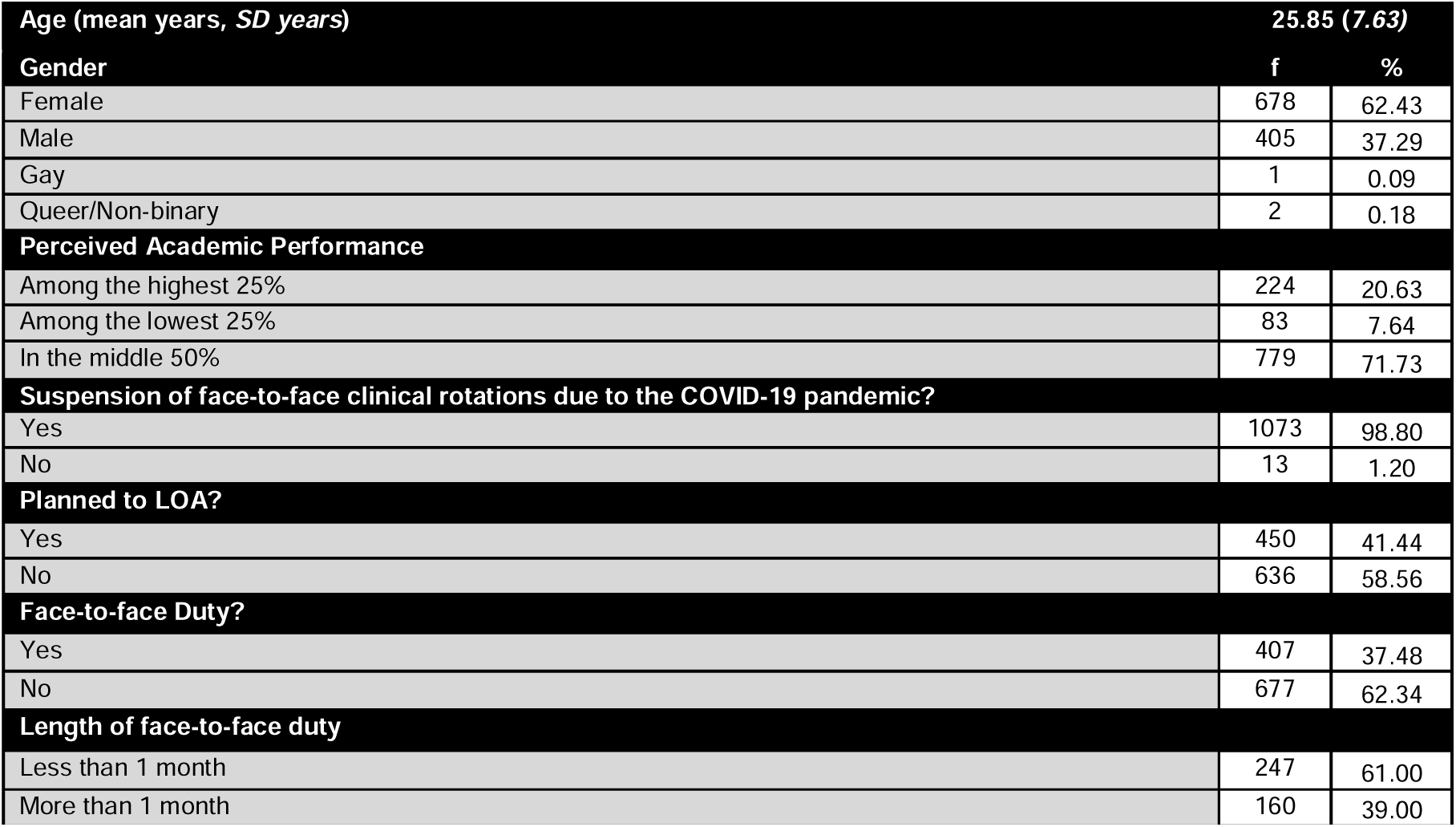
RESPONDENT PERSONAL AND LEARNING PROFILES (n=1086)

Table 2 illustrates the medical academic institutions of the respondents. The strength of this survey was being able to include the experiences and perception of the students enrolled in the medical schools found all over the country in different regions namely: Region 1, Region 2, Region 3, Region 4A, MIMAROPA, Region 5, Region 6, Region 7, Region 8, Region 9, Region 10, Region 11, and NCR.

**Table 2.**
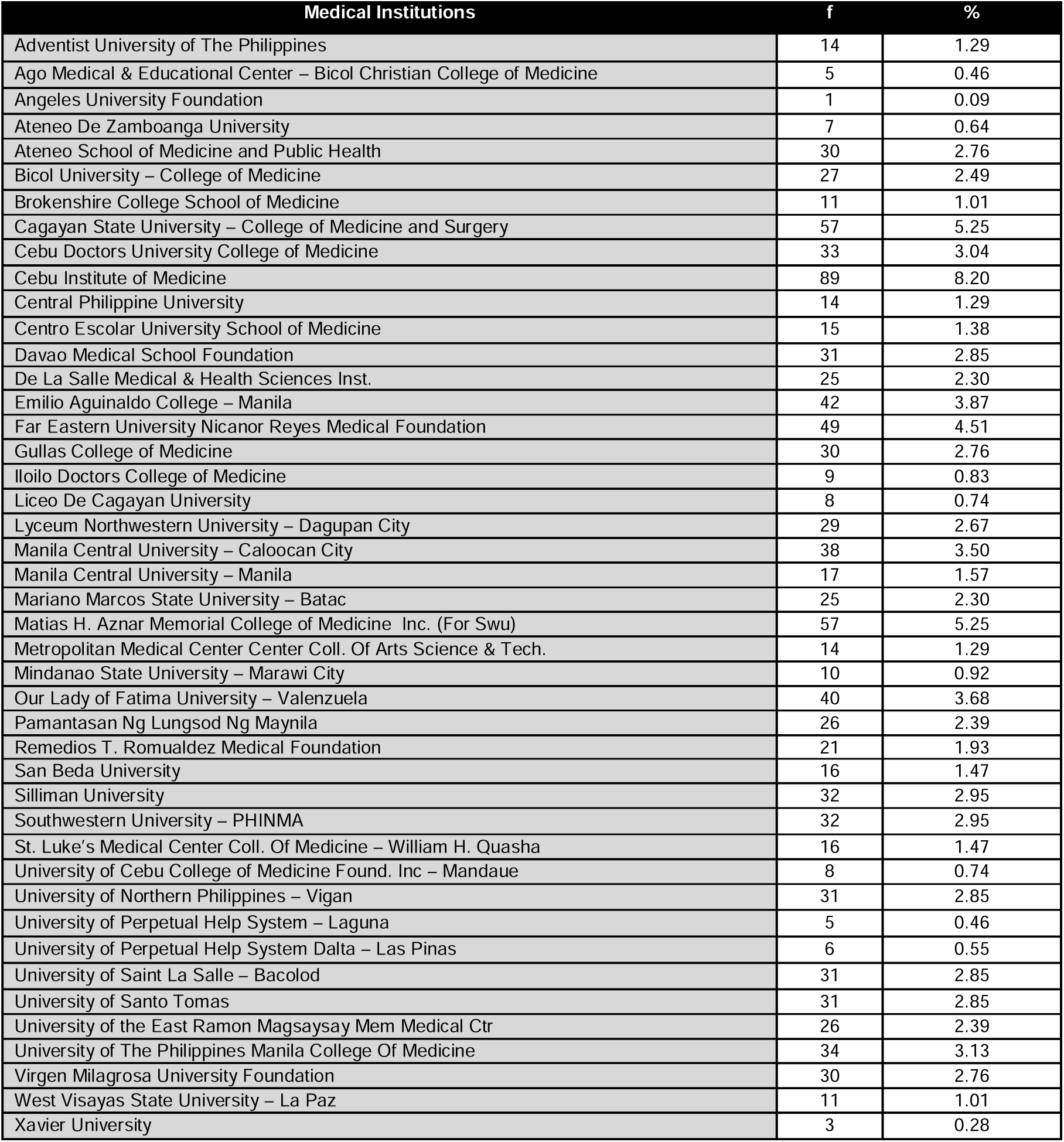
MEDICAL INSTITUTION ENROLLED (n=1086)

Table 3 enlists the most common online activities the senior medical students engaged during the pandemic. The combination of online/pre-recorded lectures, video-making, online fora / webinars / conferences recorded the highest frequency (74.31%). This is followed by online/pre-recorded lectures, online fora / webinars / conferences, case report with 12.25%.

**Table 3.**
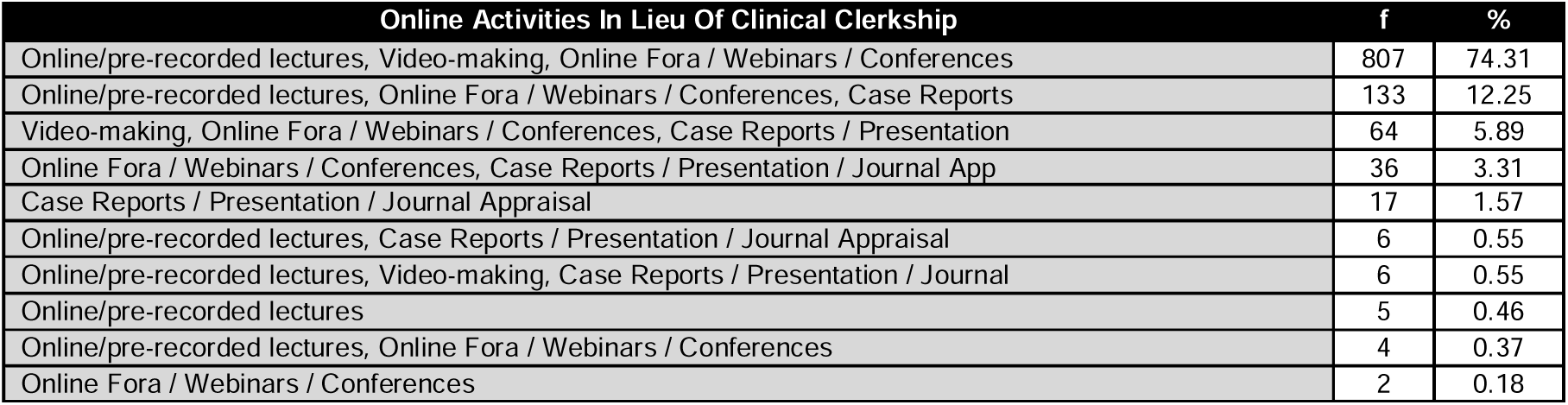
ONLINE-RELATED ACTIVITIES (n=1086)

In the area of assessment, Table 4 reveals that the students claimed that Objective Structured Clinical Examination (OSCE), Online Journal Reports, and Case Studies (41.62%) were most commonly used during this pandemic. This is followed by the assessment techniques using Online Journal Reports and Case Presentations, Situational Analysis, and Reflection at the rate of 21.73%.

**Table 4.**
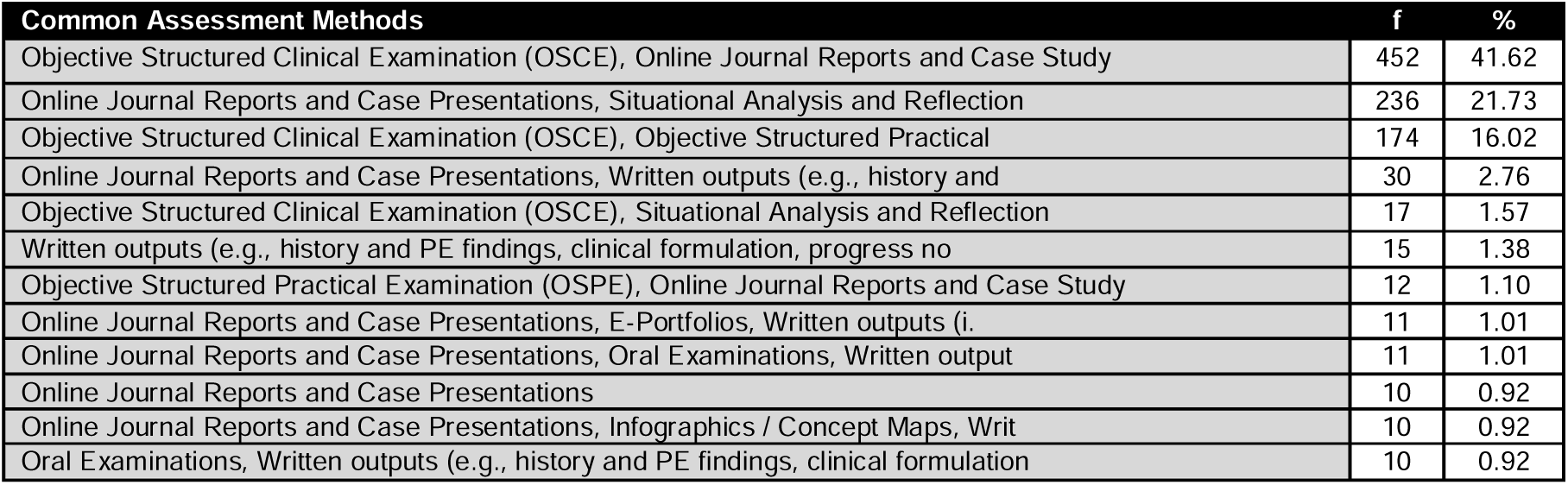
MOST COMMON ASSESSMENT METHODS USED (n=1086)

Table 5 depicts the frequently used learning management system platforms in the delivery of learning during this time of pandemic. It can be validated that Google classroom and Blackboard got the highest frequency (26.15%), followed by the use of Google classroom, Canvas, Blackboard at 7.83%. Likewise, it can be noticed that the Zoom platform was also commonly used in the delivery of lessons.

**Table 5.**
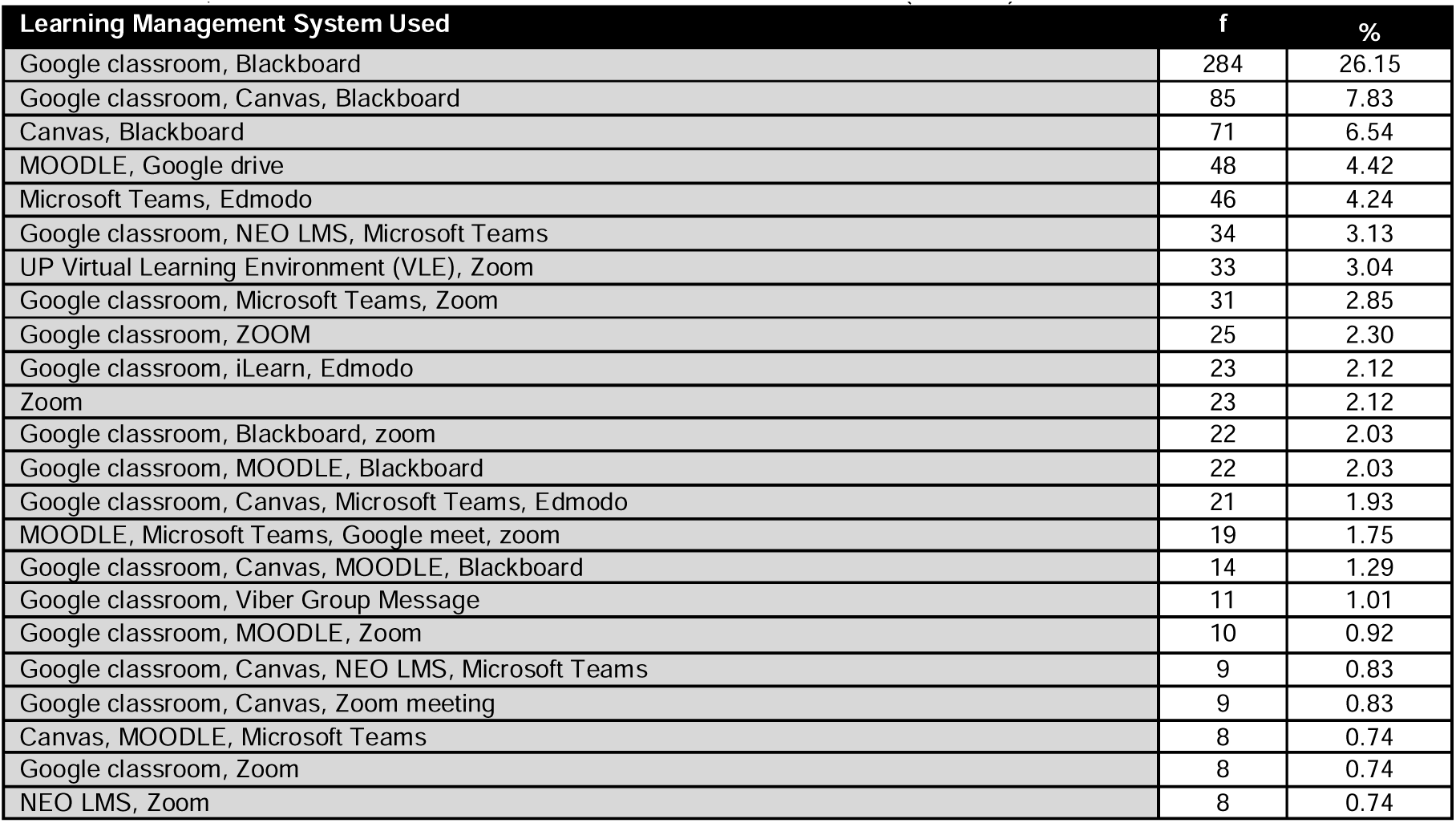
FREQUENTLY USED LEARNING MANAGEMENT SYSTEM (n=1086)

Table 6 outlines the perception of the students when asked about the outcomes of the online modality in learning. In terms of their development of skills in preparation for actual or face-to-face patient interaction, respondents generally indicated an unsure response with more than half (55.62%) of these medical learners. This is followed by their claims that they are not convinced that the online delivery of the different lessons helped them develop the necessary skills. Likewise, the majority of the respondents (79.56%) also perceived to have no confidence in assisting in hospital functions. This also showed in their negative response (44.48%) when asked about their mastery in performing physical examination (PE) and being able to identify the normal and abnormal findings with less supervision.

**Table 6.**
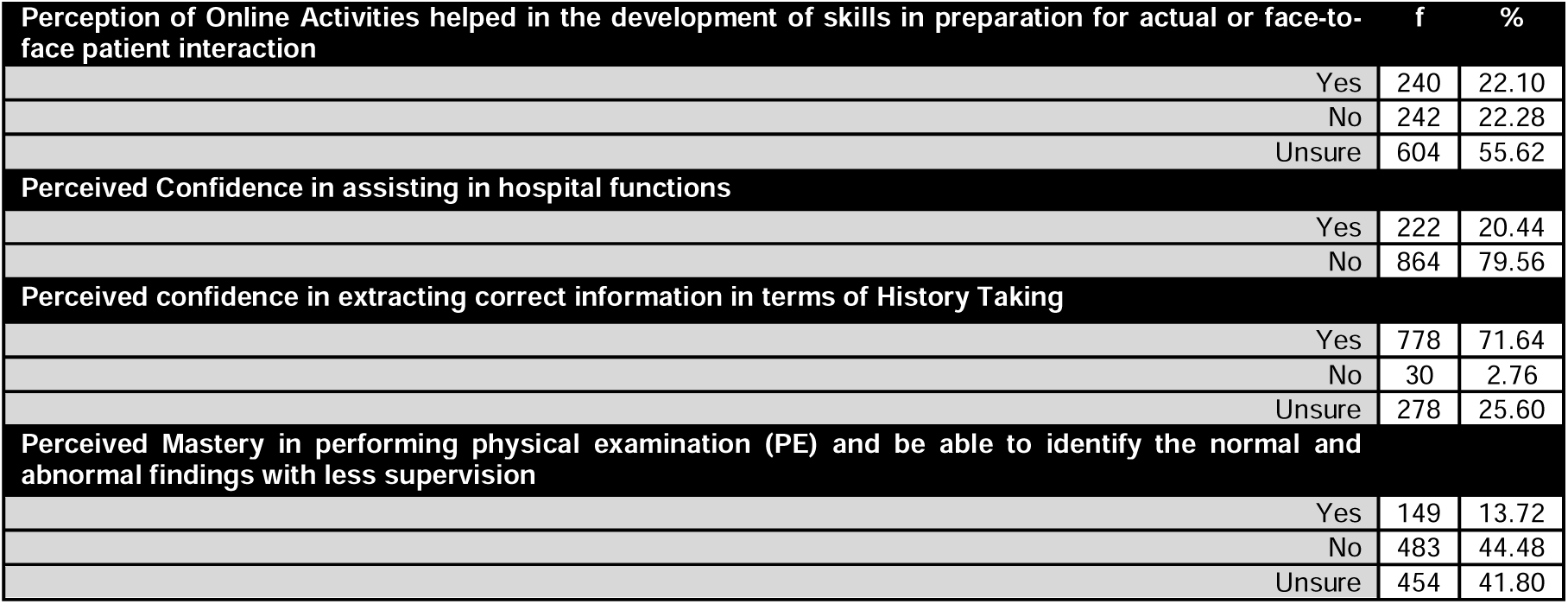
PERCEPTION OF OUTCOMES FROM ONLINE ACTIVITIES (n=1086)

### Section 2: Extent of readiness as an impact of online clinical training on the developmental areas

Table 7 shows the extent of perceived readiness of the students in terms of general clinical skills. Overall, it can be deduced that they rated this domain of readiness as “moderate” (2.92). While the respondents claimed to be “ready” for “Obtaining a complete and concise clinical history” (3.76); however, they rated “less ready” for the items on “responding effectively during emergencies, securing the airway, breathing, and circulation of the patient” (2.26) and “dealing with patients with psychiatric or psychological problems (e.g., substance abuse, psychosis) and those with cognitive impairments (e.g., dementia)” (2.33).

**Table 7.**
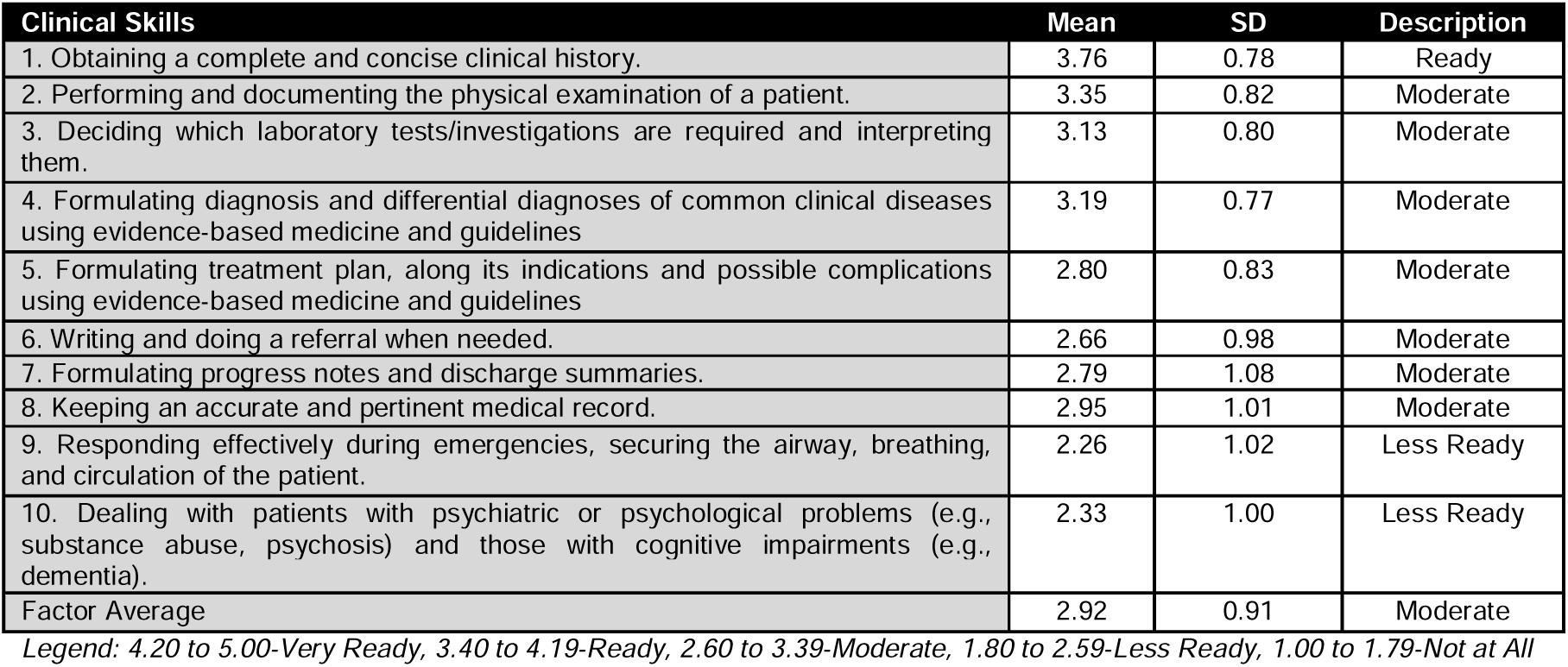
CLINICAL SKILLS (n=1086)

**Table 8.**
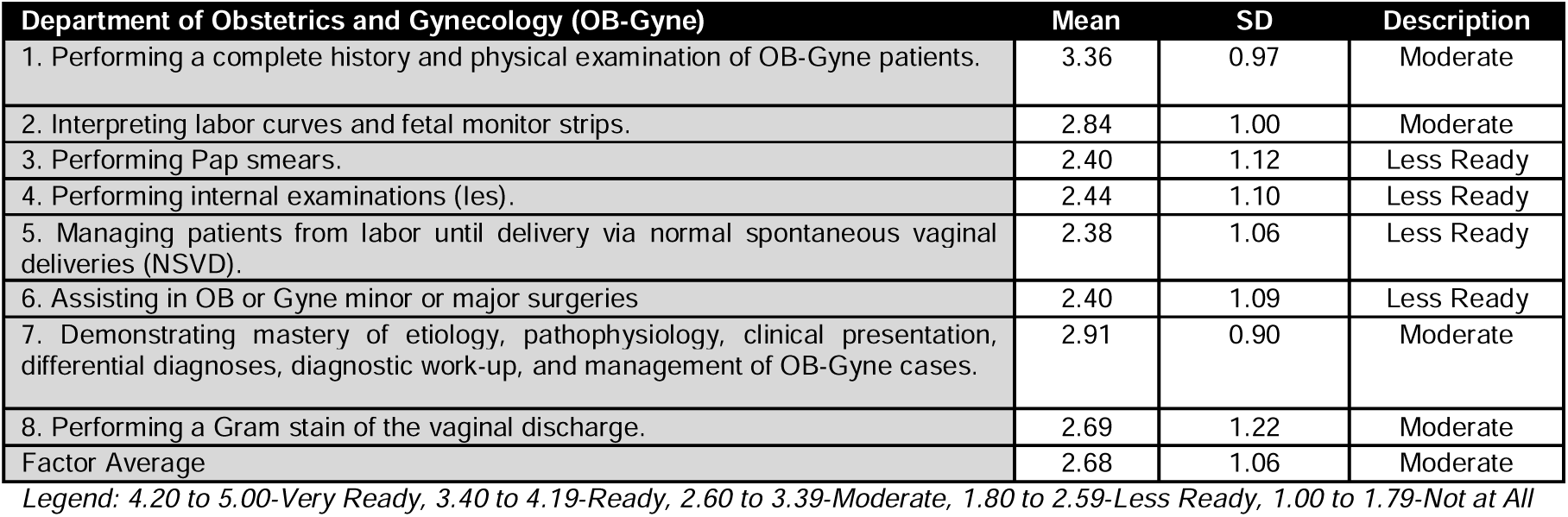
CLINICAL SKILLS IN OBSTETRICS AND GYNECOLOGY

In the area of OB-Gyne, the respondents generally rated their skills acquired from online learning as “moderate” (2.68). This can be attributed to the fact that “performing Pap smears”, “performing internal examinations”, “managing patients from labor until delivery via normal spontaneous vaginal deliveries (NSVD)” and “Assisting in OB or Gyne minor or major surgeries” were the skills they perceived to be “less ready” about.

In terms of surgery, the respondents also rated this domain of readiness as “moderate” (2.68). In fact, it can be verified from Table 9 that “Performing incision” (2.19) was a skill they perceived to be “less ready.”

**Table 9.**
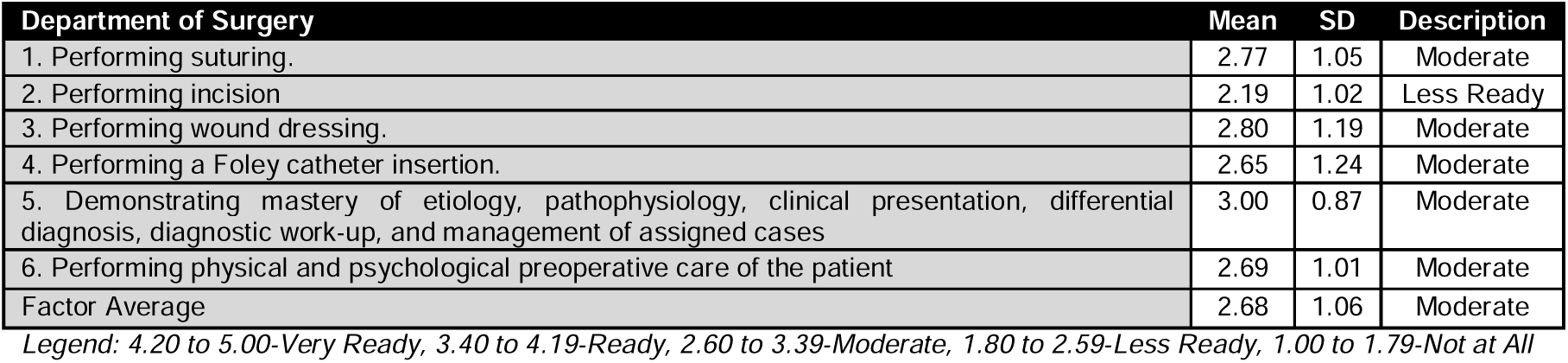
CLINICAL SKILLS IN SURGERY

Table 10 shows the perceptions of the students who claimed that they were only moderately prepared for the different tasks associated with internal medicine. It can be validated that “performing an NGT insertion” (2.50) and “Performing urethral catheterization” (2.58) were rated the least in this domain.

**Table 10.**
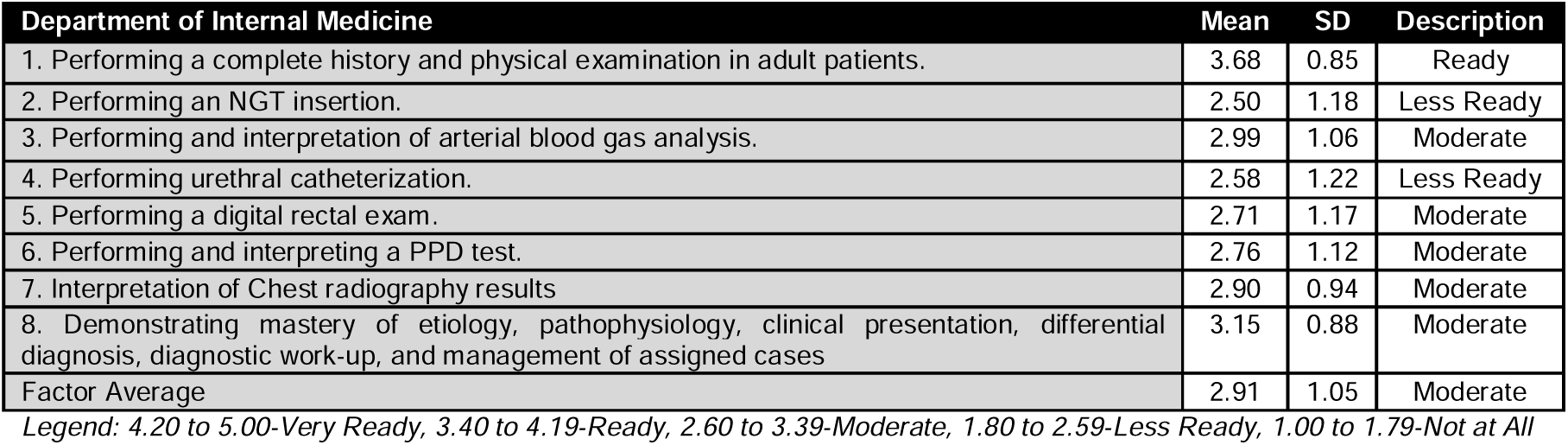
CLINICAL SKILLS IN INTERNAL MEDICINE

**Table 11.**
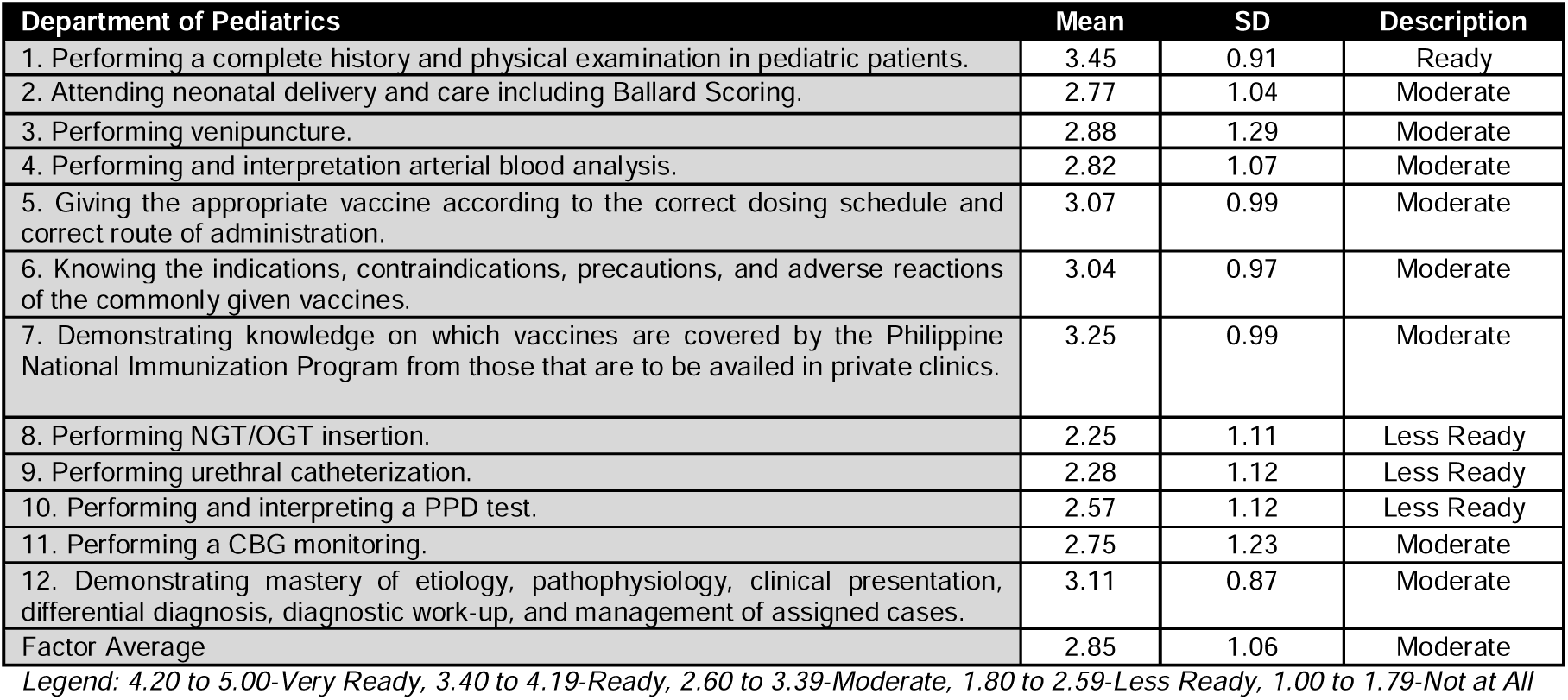
CLINICAL SKILLS FOR PEDIATRICS

In terms of pediatrics-related skills, the students rated “ready” on the item of “performing a complete history and physical examination in pediatric patients” (3.45); however, items on “performing NGT/OGT insertion”, “performing urethral catheterization”, and “performing and interpreting a PPD test” were rated as “less ready” only.

Table 12 shows that generally, the students perceived a “moderate” level (3.26) of readiness in terms of Family and Community Medicine-specific skill sets. It can be validated that they perceived the item on “identifying problems commonly encountered in rural medical practice” as “ready” already (3.40). But, the items on “doing online consultations through Telemedicine’’ and “formulating and managing team approach in implementing health programs’’ were the least rated competencies in this domain.

**Table 12.**
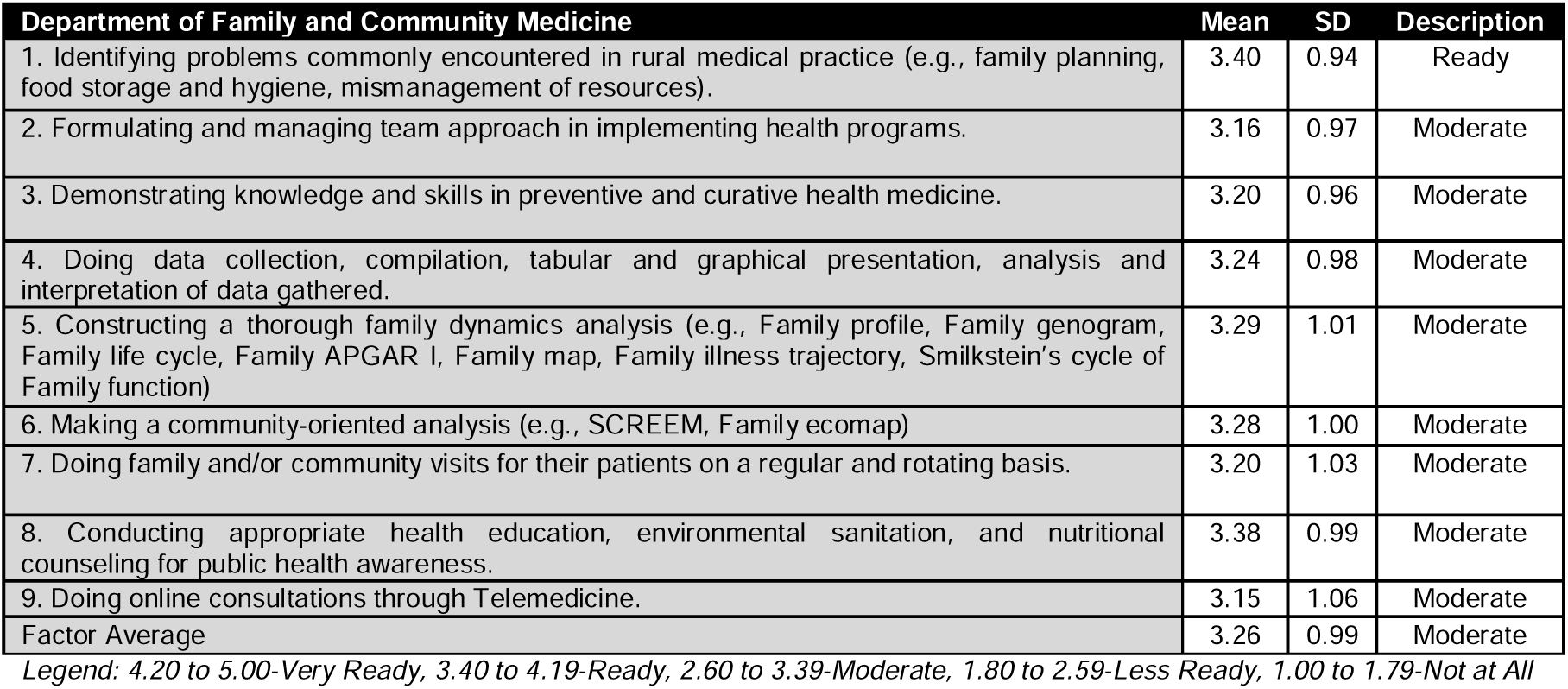
CLINICAL SKILLS FOR FAMILY AND COMMUNITY MEDICINE

As to the patient management skill sets, the respondents generally rated this domain as “ready” (3.62). This is attributable to the fact that they rated the highest perceived readiness on the aspect of “taking opportunities to promote health and prevent disease” (3.83) and “behaving with respect for patients’’ (4.15); however, the item “discussing medication, including unwanted effects, with patients’’ was rated the least (3.38) in this part of the survey.

In terms of communication and teamwork as necessary competencies at work, the students perceived that they were already “ready” (3.78) at the onset of the survey. This can be attested from their high ratings on the items for “working harmoniously with colleagues” (3.97), “maintaining good interpersonal relations with the paramedical hospital staff (nurses, medical technologist, PT/OT) and the non-medical hospital staff” (3.91) and “seeking help and advice from senior colleagues” (3.96).

Students deemed themselves “ready” in terms of the skills to understand clinical guidelines and protocols with an overall rating of 3.91.

The senior medical students also claimed that they were already “ready” (3.78) in terms of the personal development and wellbeing needed to function in their future jobs in the hospitals.

### Summary of perception on students’ level of readiness

It can be revealed from the bar graph that compliance to “Clinical Guidelines and Protocols” was the highest rated domain in terms of readiness. This is followed by “Communication and Teamwork;” and “Personal Development and Wellbeing.” Meanwhile, the skills for OB-Gyne, surgery, pediatrics, internal medical, and general clinical skills were poorly rated in terms of the level of readiness.

### Section 3: Significant difference in the readiness level of the respondents with respect to the different mandated preparatory skills

Using a One-Way ANOVA, the significance of the mean differences in the perception of readiness with respect to the different mandated preparatory skills was performed. Through the p-value approach, the ANOVA p-value recorded at 0.000 which is less than the level of significance at 0.05. This means that the null hypothesis of no significance can be highly rejected. This implies further that the differences in the extent of perception of readiness across mandated preparatory skills were statistically significant.

It can be deduced from the table that the ratings for readiness on “Clinical Guidelines and Protocol “, “Communication and Teamwork” and “Personal Development and Wellbeing” were perceived to be the highest. Since these three domains of readiness shared the same Tukey’s grouping category, then the three did not significantly differ in terms of the ratings. The readiness is then followed by patient management skills, then family and community medicine. Lastly, the Tukey’s test verified that the students rated significantly lower in terms of readiness on the items of Surgery and OB-Gyne-related skills.

## DISCUSSION

One of the immediate measures implemented by CHED to mitigate the transmission of COVID-19 was the suspension of classes, including clinical clerkship, internship, practicum, and rotation. In fact, the majority of the surveyed medical schools here in the Philippines issued a suspension of face-to-face clinical rotations in the actual hospital and community settings. To ensure the continuity of learning, a shift towards flexible learning was implemented.^6^ Most of the clinical clerkship activities are now done through online platforms, using pre-recorded lectures, video-making, online fora, webinars, and conferences. Likewise, clinical skills assessments are done online via OSCE, journal reports, and case studies.

This rapid shift to online learning may have offered advantages brought by its flexibility as to time and location, availability of an array of online tools for the delivery of lectures and activities, and the opportunity to give and receive immediate feedback.^7^ It is also said to enhance creativity and adaptability among students;^8^ however, despite this new learning approach, most medical students surveyed were unsure if such online activities aided in developing and enhancing their clinical skills; thus, they were not confident enough if they were tasked to assist in the hospital setting. It was also shown that the majority of the medical students were confident in their history taking skills, since this skill was already introduced, practiced, and assessed early in the course during SGDs and clinics rotation in both second and third years; however, many students lack confidence in performing PE and interpreting findings with less supervision. This goes to show that although online learning is the most appropriate approach at this time, certain skills in clinical clerkship may really require face-to-face delivery for students to develop mastery and gain confidence and readiness in preparation for the internship.

As a result, senior medical students may be at a disadvantage due to missed opportunities to refine essential clinical skills. There is also a lack of realism with simulation, limited clinical exposure, and absence of mentoring and supervision.^9^ This supervised student involvement in patient care has been found to contribute significantly in enhancing students’ cognitive knowledge and higher level application of clinical skills,^10^ and in turn allowing an adequate assessment of the learned skill and other competencies by their mentors. The ability to communicate with actual patients and perform physical examinations are also vital for learning and developing a good clinical eye and thought process.^11^ Indeed, patient contact provides a unique learning experience for those in the health professions, especially the medical students. The new online platform for clinical clerkship training, where patient contact is either limited or restricted due to the COVID-19 pandemic, has posed difficulties to students in honing certain patient care competencies,^8^ affecting their confidence level in performing certain skills or procedures.

In the institution of the researchers (CIM), senior medical clerks have stuffed toys as their patients and use of available materials at home. In a published 2021 paper by clinical clerks of Centro Escolar University, Manila, Philippines, they detailed their experiences and improvisations to replicate some skills. For instance, they simulated episiorrhaphy on a chicken to mimic the reproductive organ of a postpartum patient.^12^

Section 2 gauges the perceived readiness of the respondents in the different developmental areas. The results have distinguished the degree of readiness on both hard and soft skills. Hard skills are job-specific skills which are teachable abilities and can be measured through examinations.^13^ In this study, these are accounted for and itemized per department under Clinical Skills (Table 7), Obstetrics and Gynecology (Table 8), Surgery (Table 9), Internal Medicine (Table 10), Pediatrics (Table 11), and Family & Community Medicine (Table 12) where each department was rated moderately ready. On the other hand, soft skills are non-technical skills that define their relationship with others, life, and work. They are also the hardest skills to develop. Also called people’s skills, these can encompass communication abilities, language skills, empathy, time management, teamwork, and leadership traits.^13^ In this study, these skills are listed under Patient Management Skills (Table 13), Communication and Teamwork (Table 14), Understanding Clinical Guidelines and Protocols (Table 15), and Personal Development and Well-being (Table 16), each of which was rated ready.

**Table 13.**
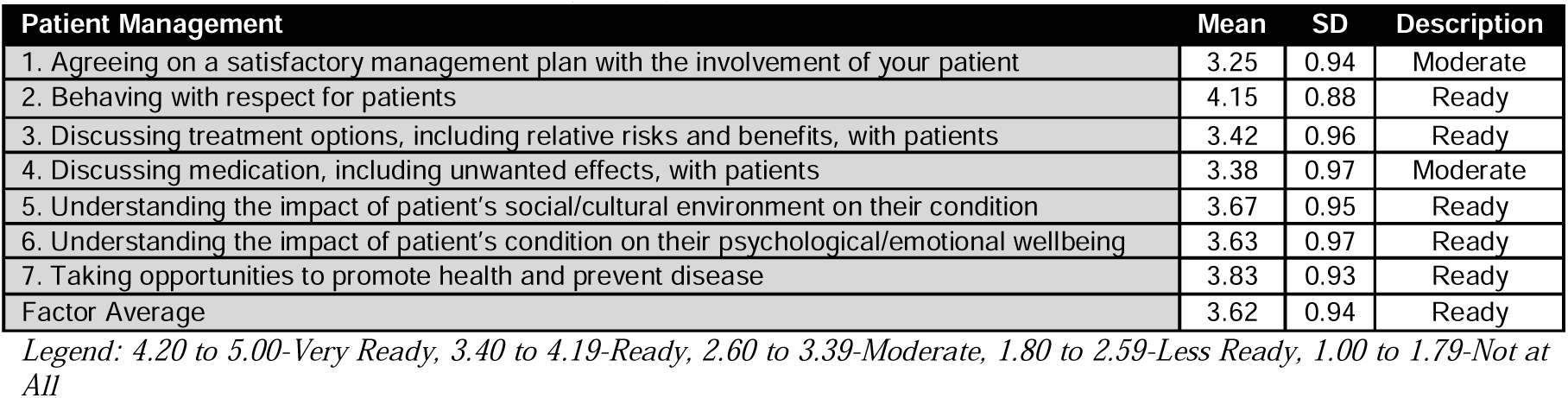
PATIENT MANAGEMENT SKILLS (n=1086)

**Table 14.**
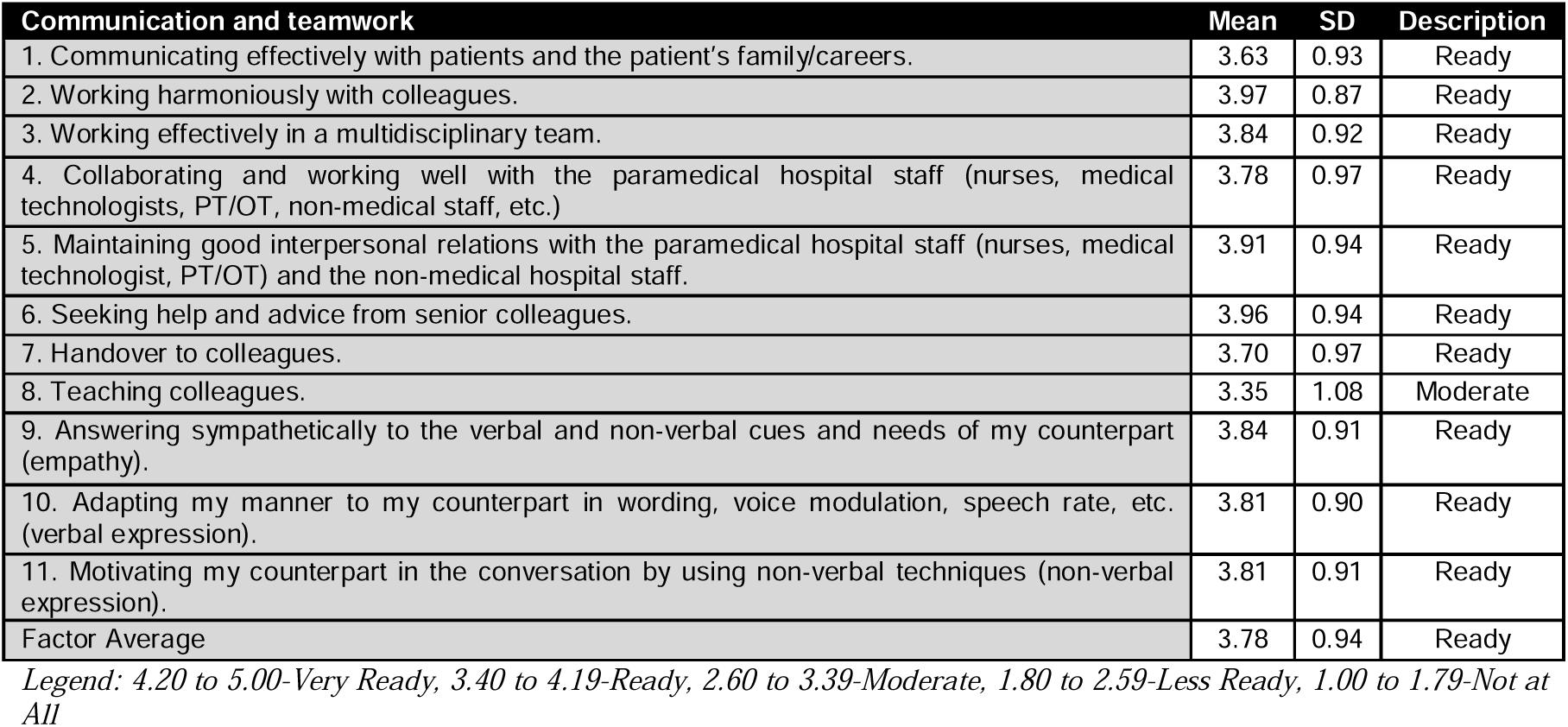
COMMUNICATION AND TEAMWORK (n=1086)

**Table 15.**
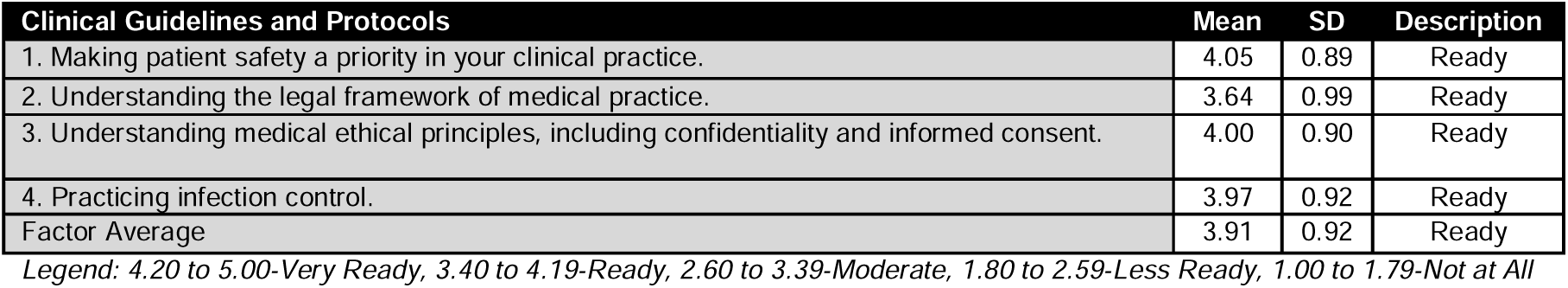
UNDERSTANDING CLINICAL GUIDELINES AND PROTOCOLS (n=1086)

**Table 16.**
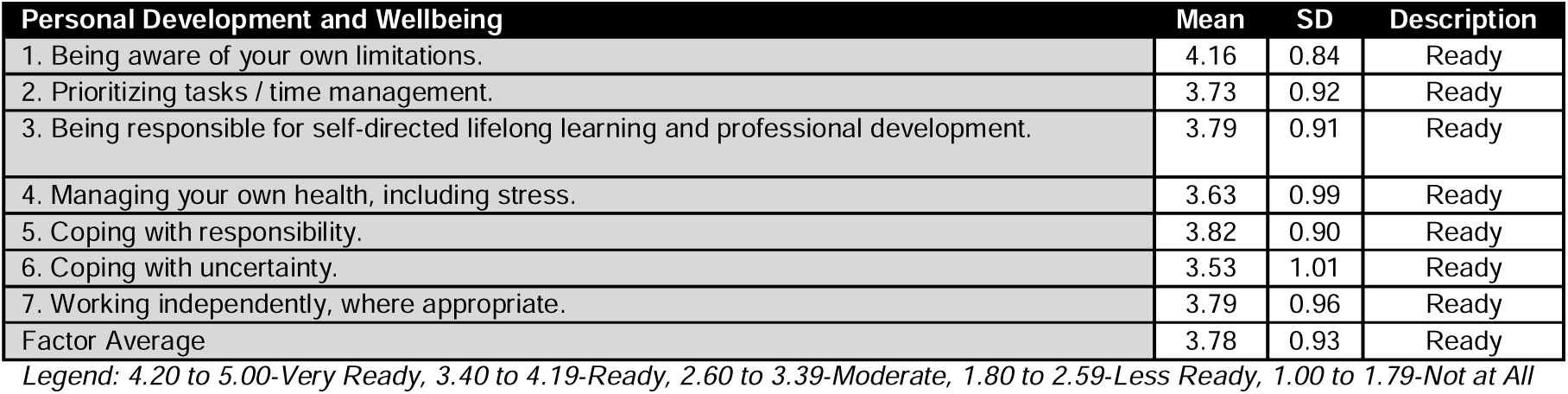
PERSONAL DEVELOPMENT AND WELLBEING (n=1086)

**Table 17.**
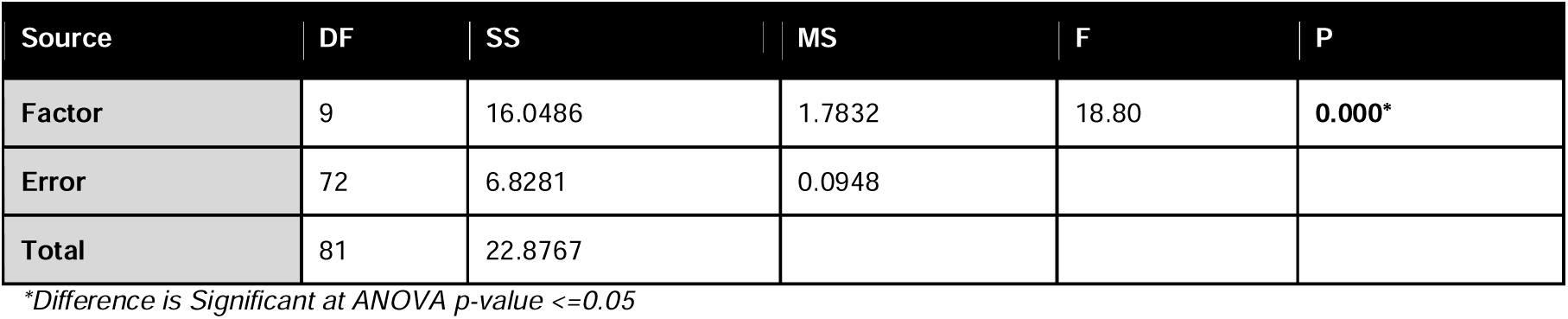
ONE WAY ANALYSIS OF VARIANCE (ANOVA): Clinical Ski, Department o, Department o, …

**Table 18.**
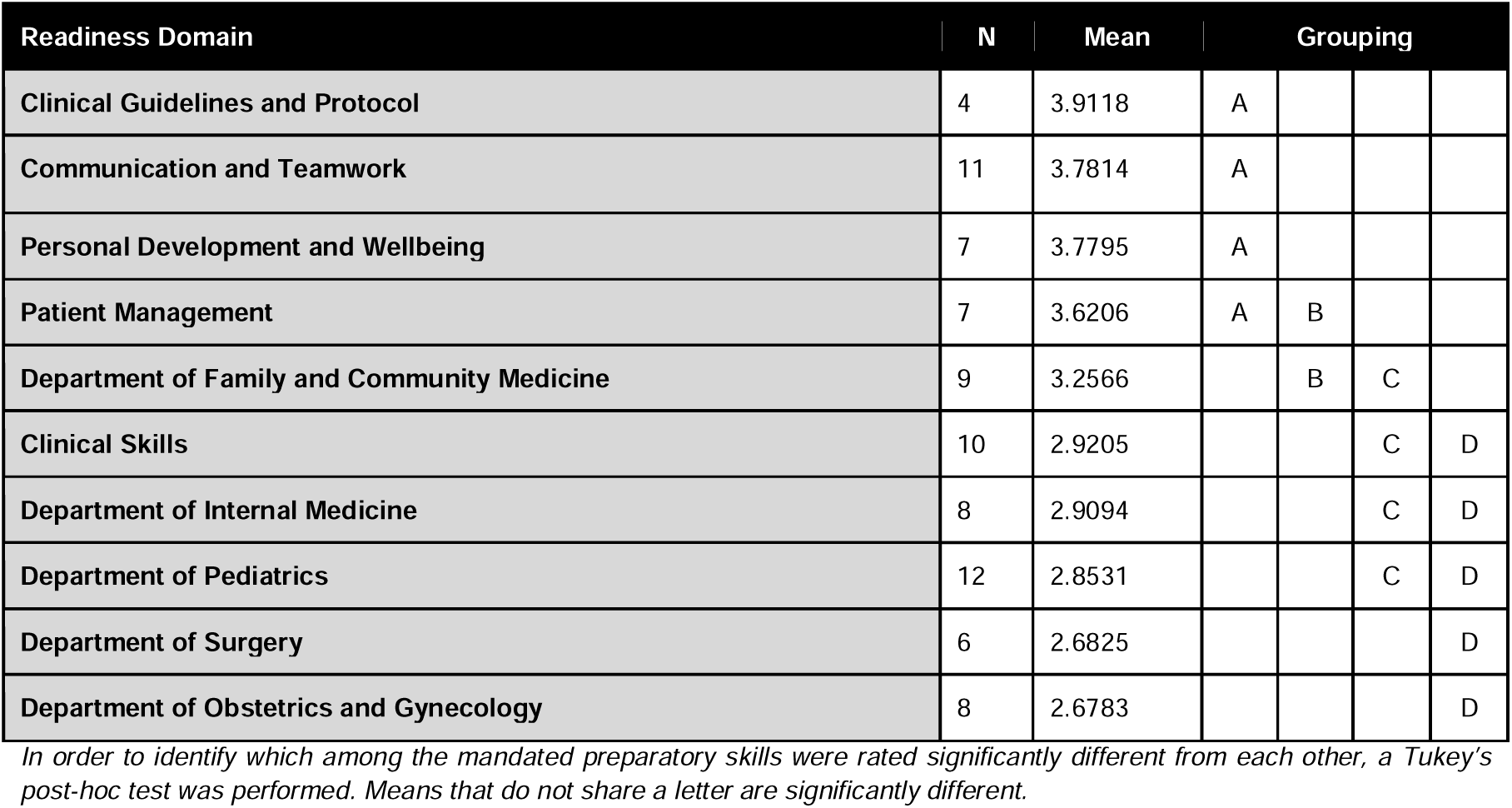
POST-HOC ANALYSIS FOR GROUPING INFORMATION USING TUKEY METHOD

Soft skills describe an individual’s emotional intelligence quotient (EQ).^13^ These can be assessed by standardized tests like Bar-On EQ-1 2.0 among others;^14^ hence, it is noteworthy that this study may not accurately measure the Emotional Intelligence (EI) of the respondents. Despite this, the results identified how the senior medical clerks deemed themselves more ready in all aspects of the soft skills than the hard skills (Figure 1). The negative impact of online learning on mastering clinical hard skills is supported by several literature. A national cross-sectional survey done to 2721 medical students from the United Kingdom (UK) showed how 75.99% of their respondents felt that online teaching had not successfully replaced clinical teaching via direct patient contact; 82.71% felt that online learning cannot equip them with practical clinical skills.^15^ Surgical and procedural (psychomotor) skills are learned skills. Dexterity (motor) alone does not equate to skill whereas skill is intelligently (psycho) applied manipulation. Assessing these skills involves measuring two tangible components: (1) the information that the trainee is using to learn, and (2) the aspect most commonly reported in the literature, the trainee’s performance, assessed through observation of the trainee. If a trainee successfully ‘shows how’ or ‘does’ an operation or procedure, this at least proves that the knowledge to perform the procedure was in store.^10^ The results of the study of the researchers paralleled the findings of another study from the UK which surveyed 440 final year medical students. It showed that disruptions to student internship had the biggest effect on their confidence and preparedness where 59.3% of students felt less prepared while 22.7% felt less confident.^16^ The perceived clinical readiness of the medical clerks can affect their confidence once there will be a shift to face-to-face patient interactions. A recent study involving 179 medical students in Singapore showed that approximately one-third did not wish to return to the clinical setting as they appear concerned about introducing possible risks to their patients as they are “not trained”.^17^ The third section depicts the significance of the mean difference in senior medical students’ perceptions following the different preparatory skills mandated by CHED. The areas where they are most confident relate to aspects of “Clinical Guidelines and Protocol,” “Communication and Teamwork,” and “Personal Development and Wellbeing.” This may be a reflection that indeed the senior clerks have honed their skills in conveying information, both in written and in oral formats utilizing different types of resources, as well as effectively working as a team with co-students, faculty staff, and other professionals in managing assigned cases and projects. In addition, this confirms their mastery in pursuing lifelong learning and personal growth through self-directed learning, developing attitudes and values essential for a primary health care physician, as the competencies aforementioned were all part of and were integrated on the first year of medical school, set by CHED Memorandum Order No. 18 on Course Objectives.^18^

**Figure 1.**
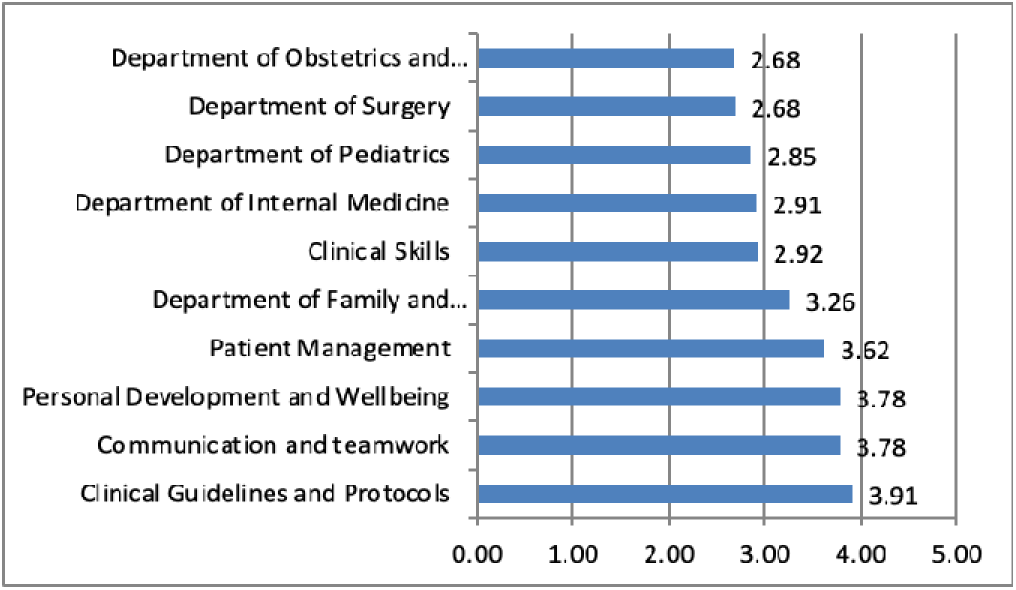
The Summary of Senior Medical Students’ Responses.

On the contrary, the respondents significantly scored lower in terms of readiness in performing skills-based practice: Surgery and OB-Gyne. This gives us an insight that the alternative means of training in this time of pandemic may have provided learning to a certain degree but is not adequate enough to replace what face-to-face training really could offer. With the transition to Telemedicine format, both aspects of medicine need further modification and training as these skills-based practices require constant exposure, experience, and repetition for enhanced skill proficiency and mastery. With these limitations brought about by the pandemic, there will be a reduction of clinical opportunities for medical students before applying to residency programs. Some medical students may miss opportunities such as a full rotation in their specialty of interest, there would also be limited exposure to certain areas of medicines and the absence of being mentored.^9^ Active contribution to patient care enhances medical student learning, but interruptions to learning have lasting effects on competitiveness for residency applications and preparedness for intern year.^19^

This study involving more than a thousand students all over the country provides information that may be used as a basis for adjustments in teaching strategies of essential skills in clinical clerkship as the entire medical educational system adapts to the new normal post-pandemic.

## CONCLUSION

The impact of COVID-19 on fourth year medical students’ online clerkship has been significant in the Philippines. The pandemic disrupted the students’ confidence and preparedness. This poses substantial issues for the learning experience and development of medical students that lead to a reduction in their clinical competence. The results of the study distinguished the degree of readiness on hard and soft skills. Hard skills are job-specific skills that are teachable abilities while soft skills are non-technical skills that define the relationship with others, life, and work.

The fourth year students from medical schools with online clerkship training felt prepared for the clinical guidelines and protocols, communication and teamwork, and personal development and well-being; however, they felt less prepared in performing the skills for OB-Gyne, surgery, pediatrics, internal medical, and general clinical skills. The results identified how the fourth year medical students deemed themselves more ready in all aspects of the soft skills than the hard skills. This showed that online clerkship in this time of pandemic may have provided learning to a certain degree but it is not enough to replace what face-to-face training could offer.

## RECOMMENDATIONS

Given the aforementioned results, the following are recommended:

### Medical School

1. Schedule students for access to on-campus facilities for skills development and simulation (e.g., skills laboratory).
2. Create opportunities for clerk-to-patient interactions (e.g., telemedicine, hospital rotations).
3. Make clinical clerks and instructors aware of the clerk’s current confidence levels to identify which areas they need to focus on improving. Let students evaluate their performance in every department (e.g., standardized questionnaire).
4. Let clinical instructors give real-time, individual feedback with regards to the clerk’s performance in terms of soft and hard skills.
5. Set a strong system in ensuring the fairness of the online summative assessment.
6. Further improve online clinical training with the use of sophisticated technologies aimed to mimic bedside assessment and clinical scenarios.
7. Allocate funds to secure and give out essential materials needed for skills practice.
8. Prioritize continuity of learning through blended learning to ensure that the medical students meet the required learning objectives and the standards set by CHED. Create and implement guidelines regarding the resumption of face-to-face classes and develop a bridging program to ease the transition from online learning to face-to-face classes.
9. Encourage students to actively participate during clinical skill training by checking on student comprehension and posing questions to maintain student enthusiasm and interest.
10. Further give all-encompassing help to the students, for example, managing the emotional well-being impacts of the pandemic and recognizing approaches to limit these adverse consequences.

### Medical Students

1. Continue developing skills through practice.
2. Utilize available facilities as well as opportunities for clinical exposure/hands-on experience.
3. Maximize the use of reliable online resources for supplementary use.
4. Assimilate further knowledge by joining webinars and medical conferences.
5. Effectively take part in the conversation by posing inquiries and reacting to them during class meetings and clinical skill training.
6. Actively participate in educating the community regarding common medical misconceptions and other health-related issues.
7. Provide input on their peers and teachers to allow every member an opportunity to ponder their insight and convey their feedback in a steady and organized manner.

### The community

1. Set clear guidelines to guarantee persistent instructive improvement of the medical students during crisis or pandemics.
2. Provide a stronger vaccination campaign in prioritizing medical students and frontline workers with clinical responsibilities.
3. Maximize resources in the surveillance and monitoring of the COVID-19 pandemic.

### Future researchers

1. Construct an extensive questionnaire with regards to assessing skills in each specific department.
2. Assess the number of hours the clinical clerks get to practice a hard skill in a specific department.
3. Determine methods used by clerks to further improve their skills outside hospital rotations and skills laboratory e.g., methods used to practice at home.
4. Maximize response rate by sending the study questionnaire directly to the medical school deans, class representatives and providing updates for completion.
5. Utilize a more systematic sampling method to capture a better representation of the population.
6. Further study on the long-term impact brought about by the absence of clinical training for several months and identify competencies that have not been fully accomplished by the medical students.

## Supporting information

IRB

## Data Availability

All data produced in the present study are available upon reasonable request to the authors

## ACKNOWLEDGMENT

Our deepest gratitude and appreciation for the guidance and support are extended to the following persons who have contributed to the fulfillment of this study:

To the Almighty, for making all things possible, giving the researchers the guidance for the fulfillment of this study.

To our research advisor, Dr. Maria Philina Pablo-Villamor, for imparting her knowledge and expertise, which meant so much for the completion of this study.

To our statistician, Sir Mark S. Borres, for imparting his expertise on statistical analysis and for his guidance in the interpretation and recommendations for the betterment of this study.

To Dr. Wilfredo M. Ypil and Dr. Ramon L. Arcadio for their suggestions and constructive criticisms.

To our dean, Dr. Martiniano C. Zanoria, for his unrelenting support, especially amid the current pandemic.

To the school deans for allowing us to gather the necessary responses from their respective schools.

To the respondents of the study, for being cooperative and allocating some of their time in answering our surveys.

## APPENDICES

## APPENDIX A. Research Instrument

### Perceived Clinical Readiness of Senior Medical Students as Outcomes of Online Clerkship in the Philippines: New Normal in Medical Education

**Dear Respondents**:

Good day!

We are third year medical students of the Cebu Institute of Medicine (CIM), conducting this survey for our study, “Perceived Clinical Readiness of Senior Medical Students as Outcomes of Online Clerkship in the Philippines: New Normal in Medical Education”. This study will determine the impact of the new online teaching platform and the alternative clinical clerkship training programs on the clinical readiness of the fourth year medical students in the Philippines.

To accomplish this endeavor, we would appreciate greatly for you to take part in this study by answering this questionnaire. It shall approximately take 15-20 minutes of your time, with data entered electronically into secure databases. Hence, we assure you that the information you provided us will be kept with utmost confidentiality and will be permanently deleted once it has served its purpose.

If you wish to continue and be part of our study, you may start answering the questions in the next few sections; however, if you do not want to participate, you may exit this Google Form at any time as your participation is entirely voluntary. We hope for your full cooperation.

Thank you very much.

Group 16, PBL-3

### I. Profiling

Age:____________

Gender:____________

Medical institution currently enrolled:___________________

Place of Residence:___________________

1. Based on your performance last academic year, your grades will fall under which of the following?
  ○ Among the highest 25%
  ○ In the middle 50%
  ○ Among the lowest 25%
2. Did your medical school suspend face-to-face clinical rotations due to the COVID-19 pandemic?
  ○ Yes
  ○ No
3. Have you ever considered taking a leave of absence?
  ○ Yes
  ○ No

### II. Online Clerkship Activities

1. Did your medical school require and/or encourage online learning while clinical rotations are being suspended? *Online learning is defined as the use of electronic technology and media to deliver, support, and enhance both learning and teaching and involves communication between learners and teachers utilizing online content. (Howlett et al., 2009) This may be synchronous (e.g., online lectures where teachers and learners have to be connected to the Internet at the same time) or asynchronous (e.g., teachers upload videos or handouts to be studied by the learners at their own time and pace).
  ○ Yes
  ○ No
2. What were the activities given online in lieu of your clinical clerkship? *Check all that applies*
  ○ Online/pre-recorded lectures
  ○ Video-making
  ○ Online Fora / Webinars / Conferences
  ○ Case Reports / Presentation / Journal Appraisal
  ○ Others (please specify):___________________
3. How were your skills evaluated or assessed by your clinical preceptor? *Check all that applies*
  ○ Objective Structured Clinical Examination (OSCE)
  ○ Objective Structured Practical Examination (OSPE)
  ○ Online Journal Reports and Case Presentations
  ○ Situational Analysis and Reflection Papers
  ○ E-Portfolios
  ○ One-on-one Return Demonstrations
  ○ Oral Examinations
  ○ Infographics / Concept Maps
  ○ Written outputs (e.g. history and PE findings, clinical formulation, progress notes, etc.)
  ○ Video Conferences
  ○ Others (please specify):___________________
4. What Learning Management System does your school utilize? *Check all that applies*
  ○ Google classroom
  ○ Canvas
  ○ NEO LMS
  ○ MOODLE
  ○ Blackboard
  ○ iLearn
  ○ Microsoft Teams
  ○ UP Virtual Learning Environment (VLE)
  ○ Others (please specify):___________________
5. Did the activities help you hone your skills in preparation for actual or face-to-face patient interaction?
  ○ Yes
  ○ No
  ○ Unsure
6. If you are asked to assist in hospitals, would you be confident doing so?
  ○ Yes
  ○ No
7. What could possibly hinder you from performing well in clinical rotations?___________________________
8. In History Taking, will you be able to extract pertinent information regarding your future patient’s case?
  ○ Yes
  ○ No
  ○ Unsure
9. Can you say you’ve mastered performing physical examination and be able to identify the normal and abnormal findings with less supervision?
  ○ Yes
  ○ No
  ○ Unsure

### III. Medical Training

For the next few questions, please base your answers on your overall learning experience. As a senior clerk and soon-to-be physician, please rate (how well or badly/ how confident) do you think you can perform the items being asked. Consider, however, how the pandemic brought impactful changes to the mode of learning, from actual patient interaction to online courses.

You may refer to this rating scale:

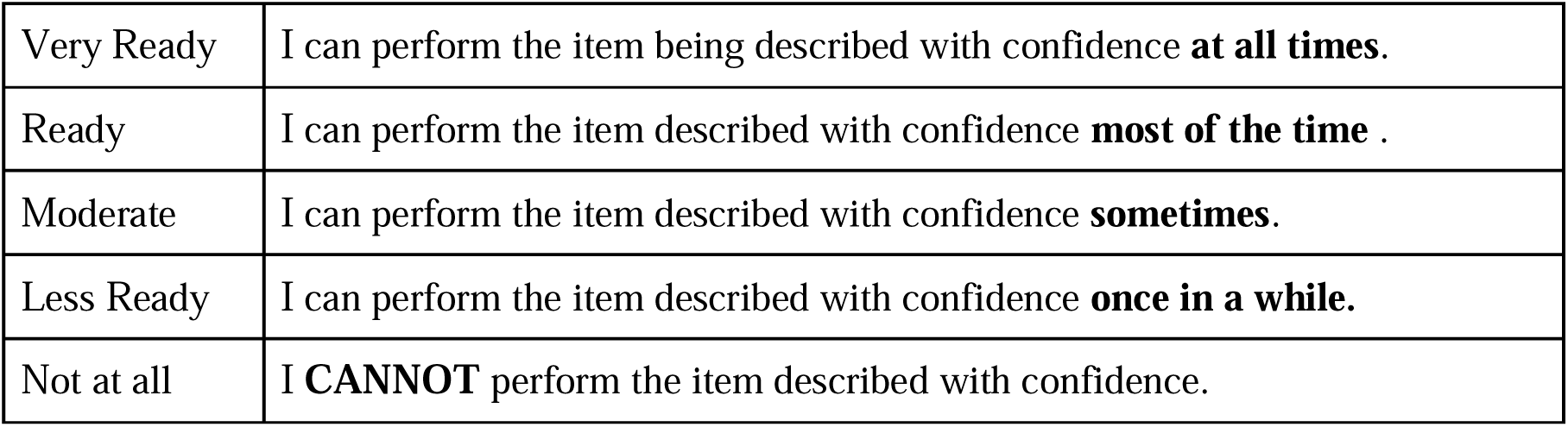

#### A. Clinical Skills

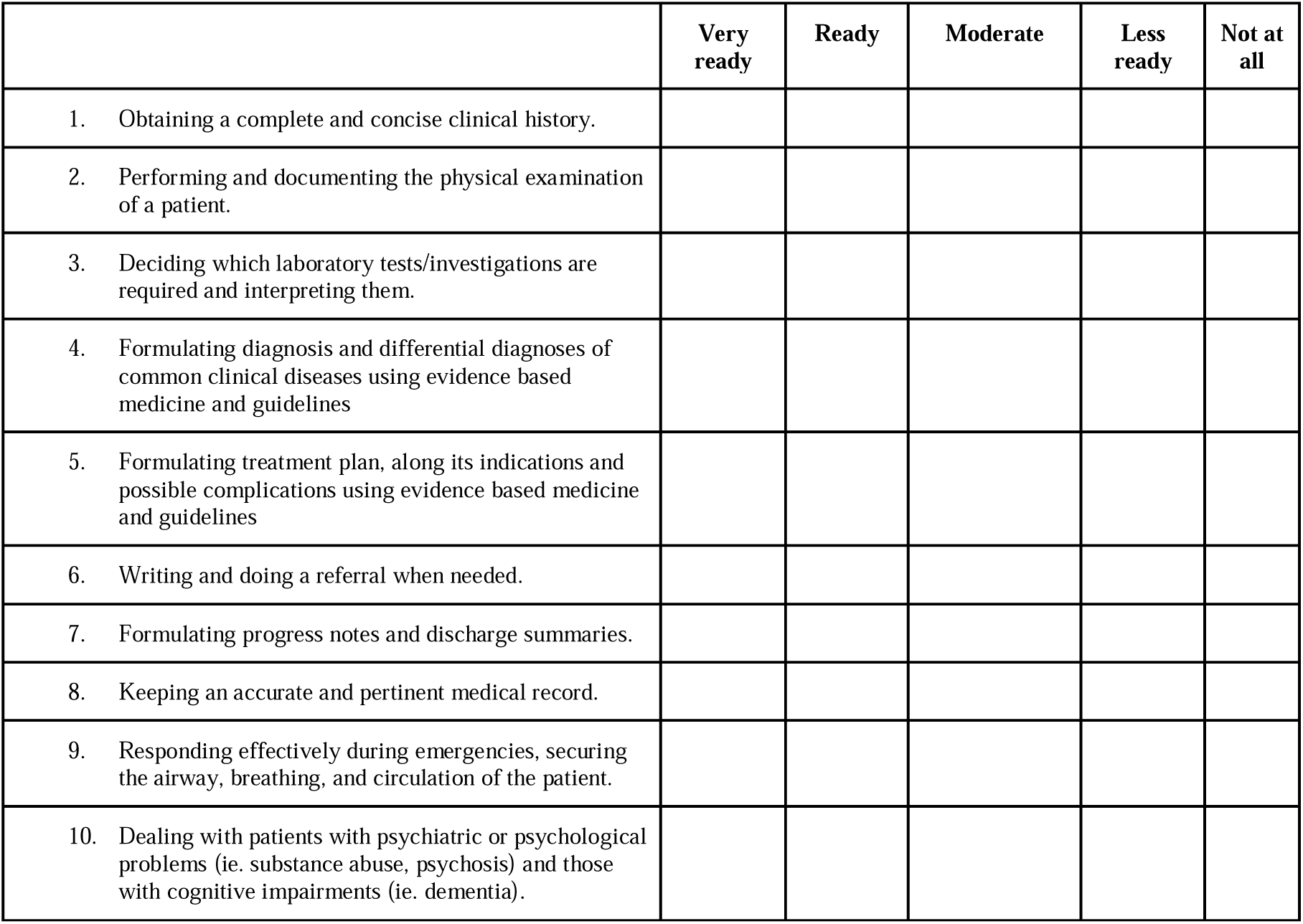

#### Department of Obstetrics and Gynecology (OB-Gyne)

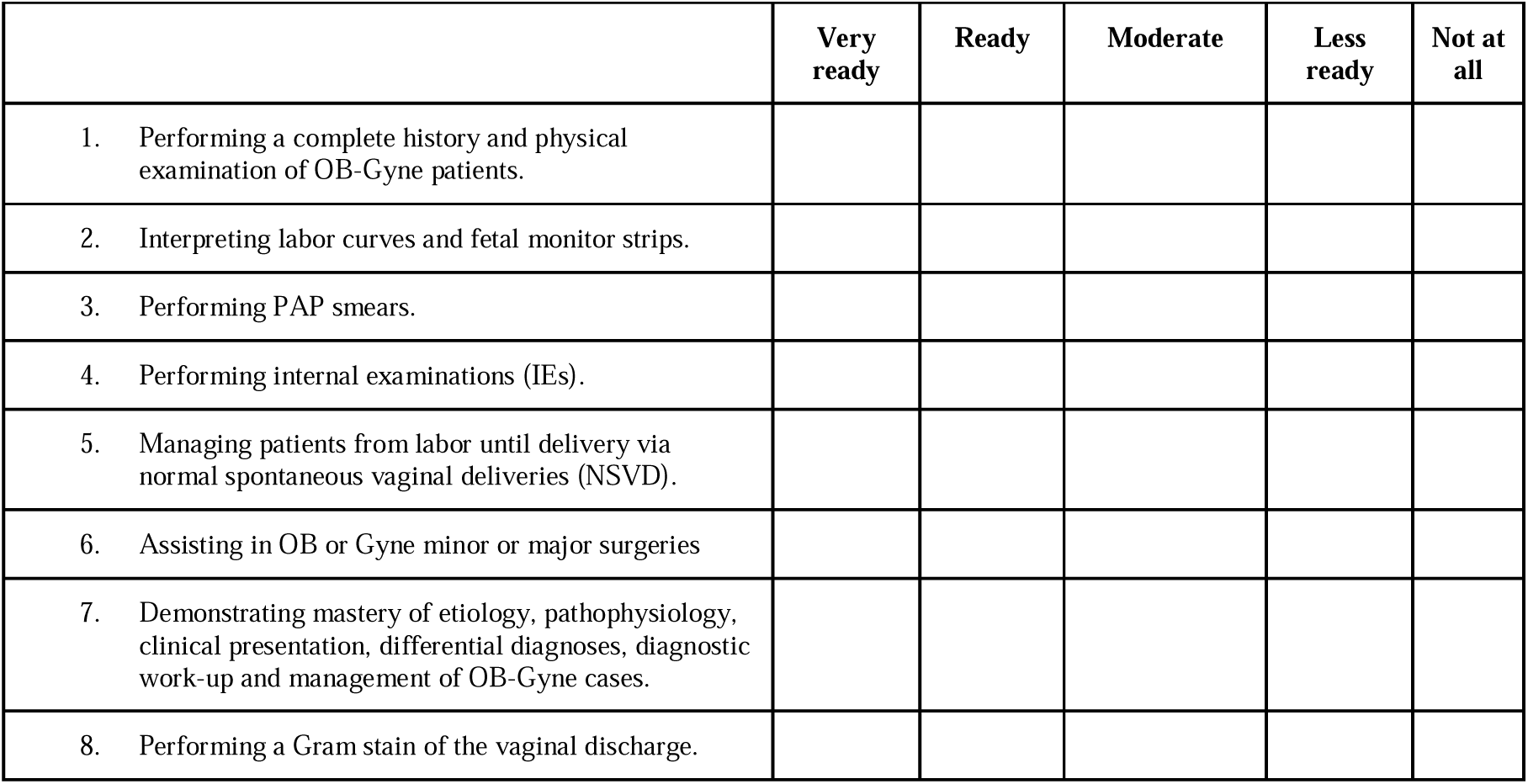

#### Department of Surgery

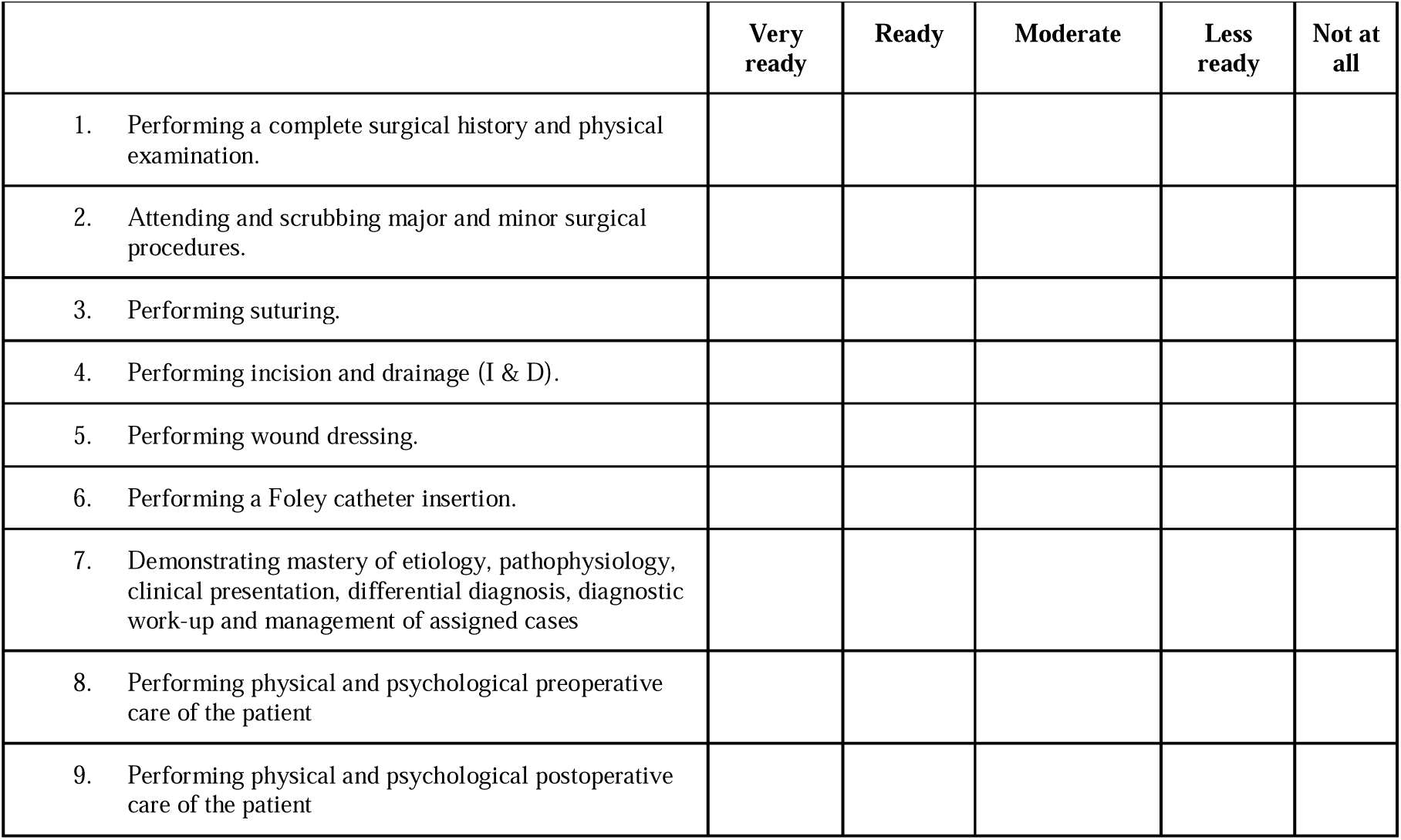

#### Department of Internal Medicine

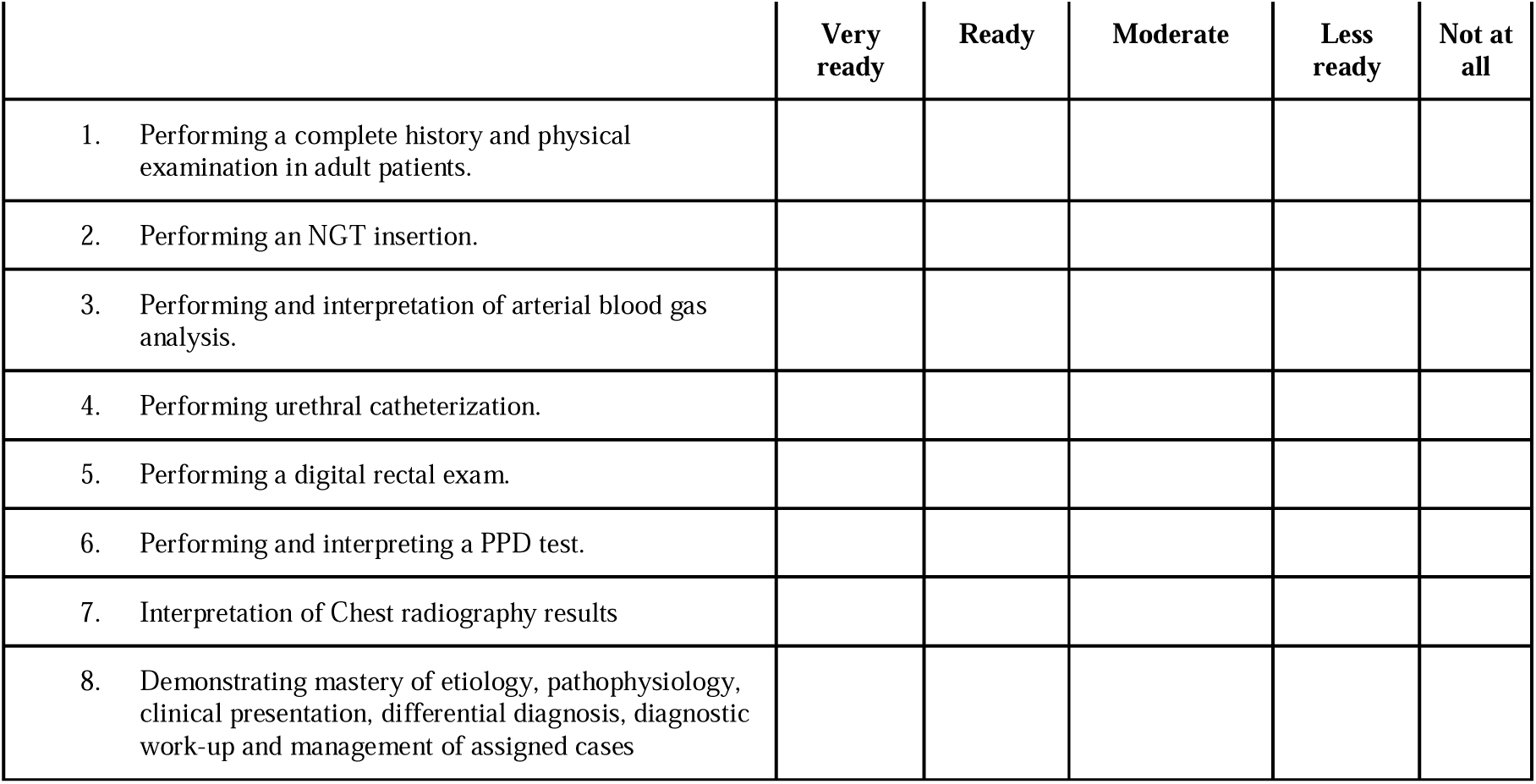

#### Department of Pediatrics

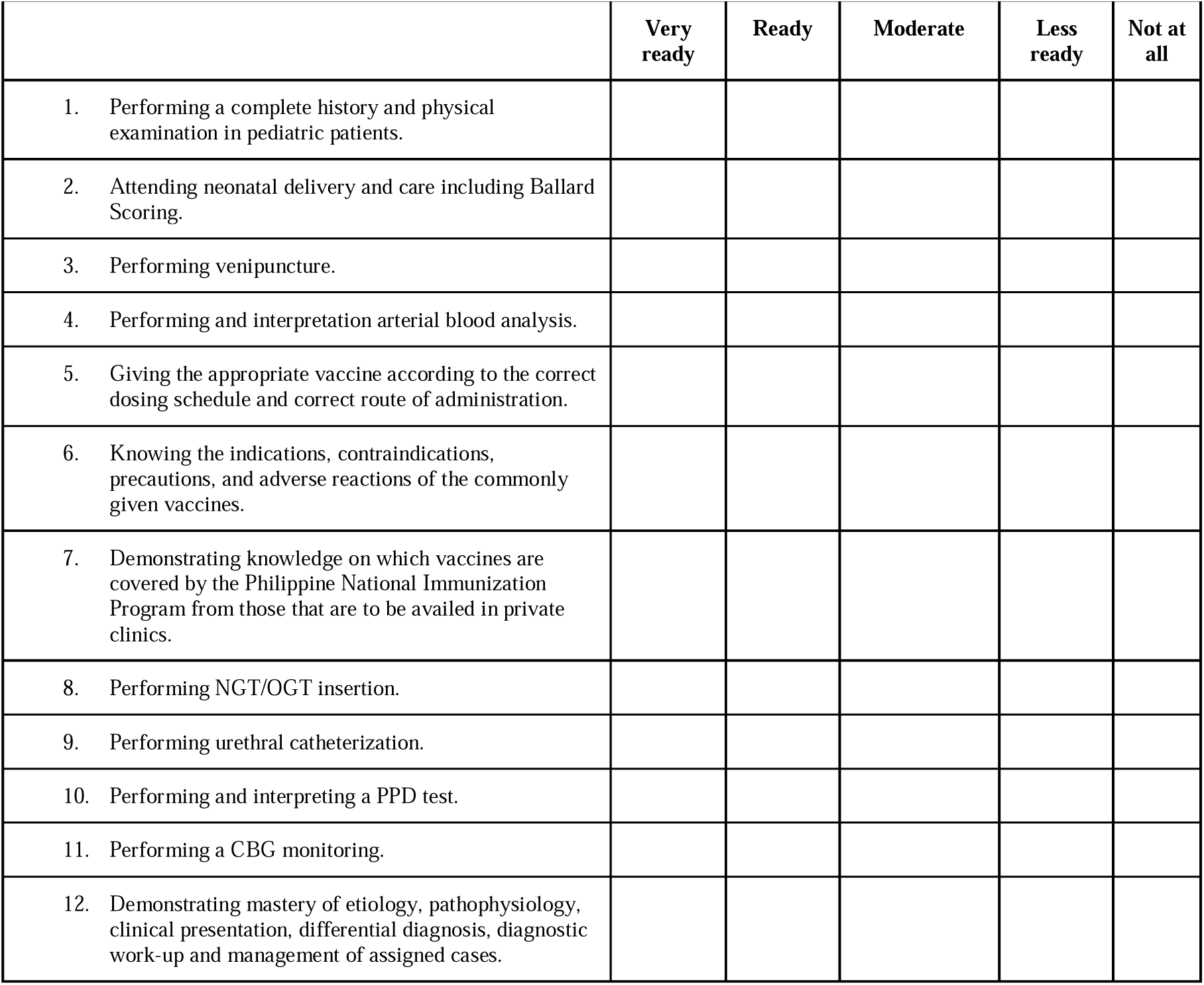

#### Department of Family and Community Medicine

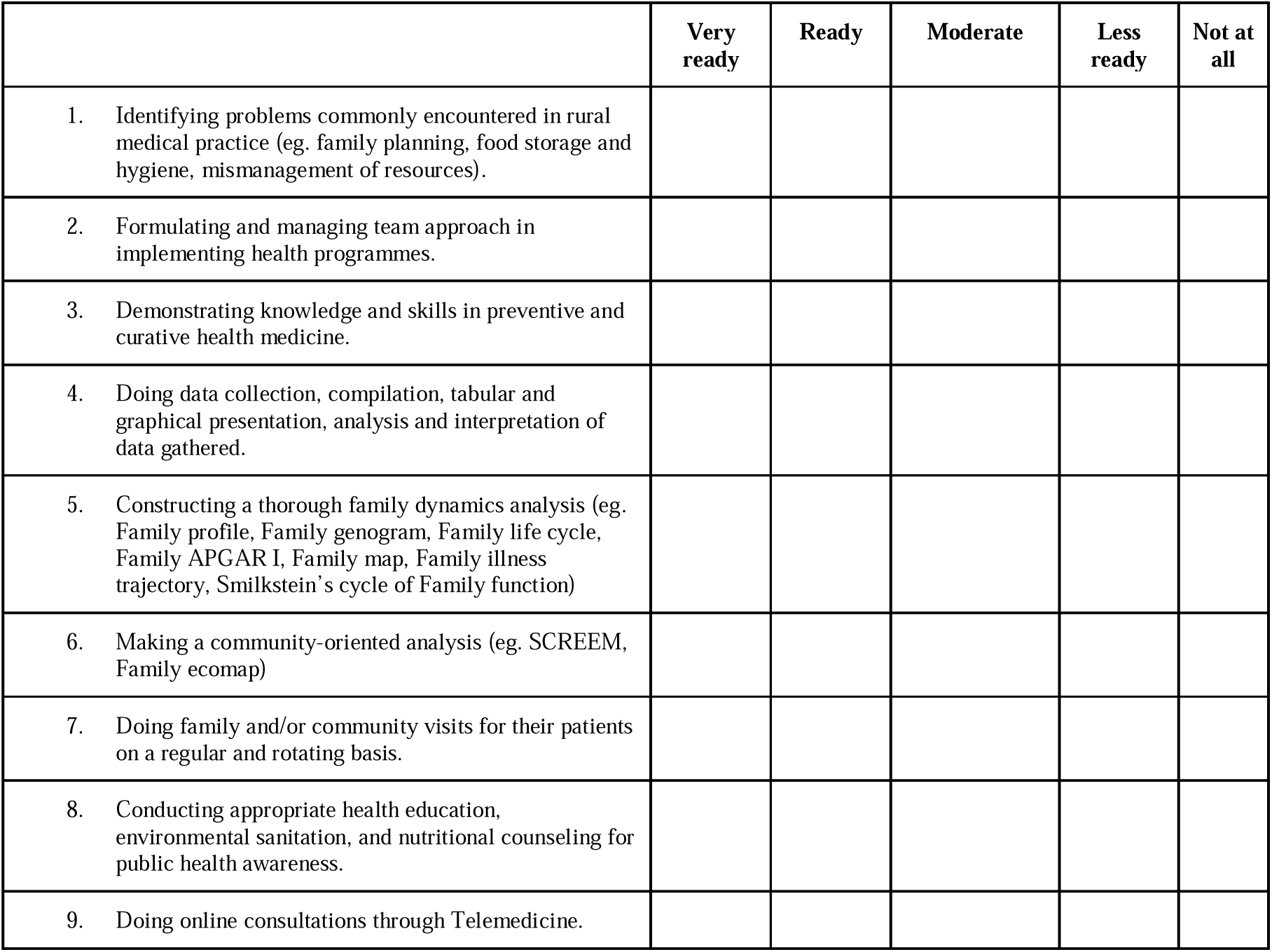

#### B. Patient Management

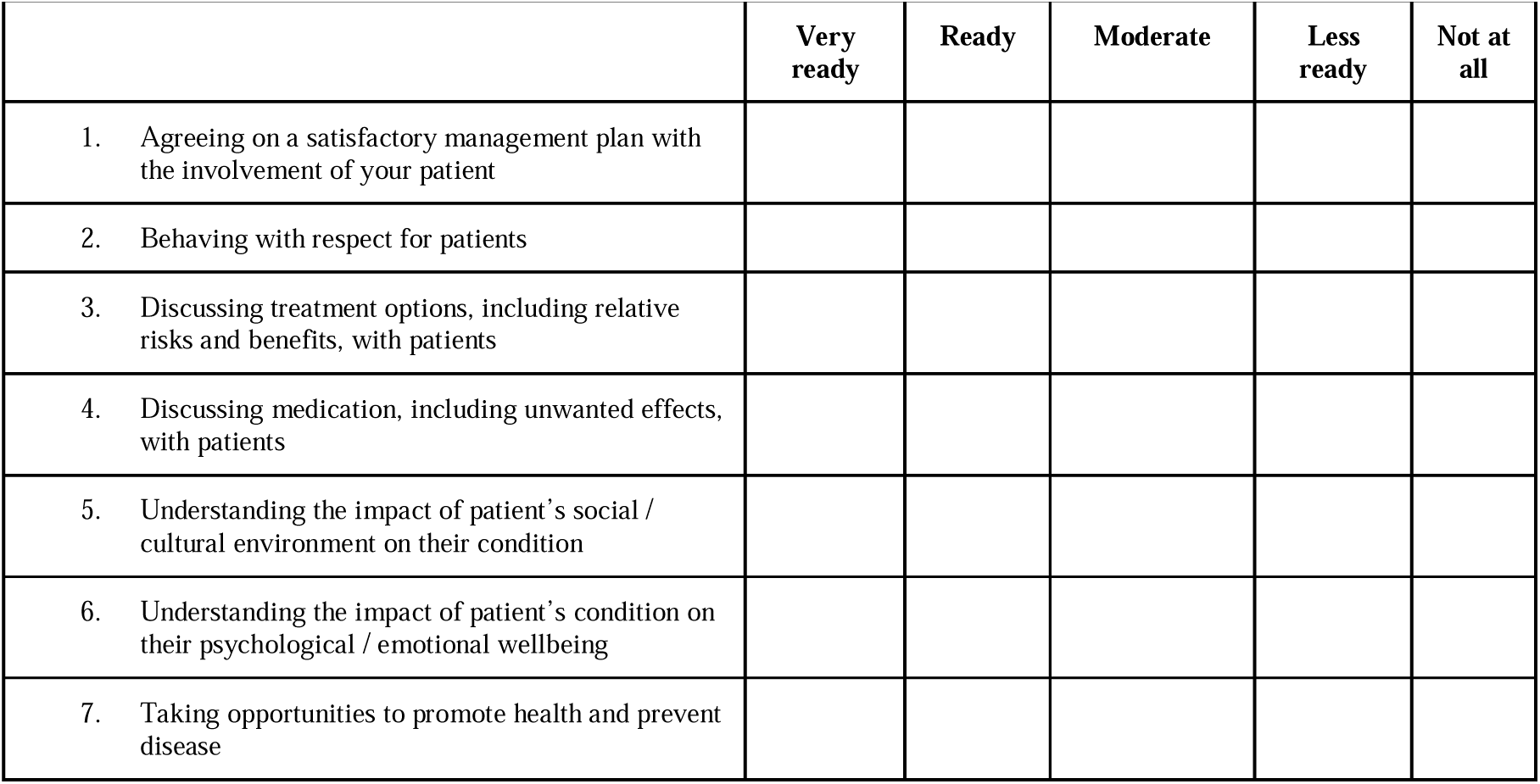

#### C. Communication and team-working

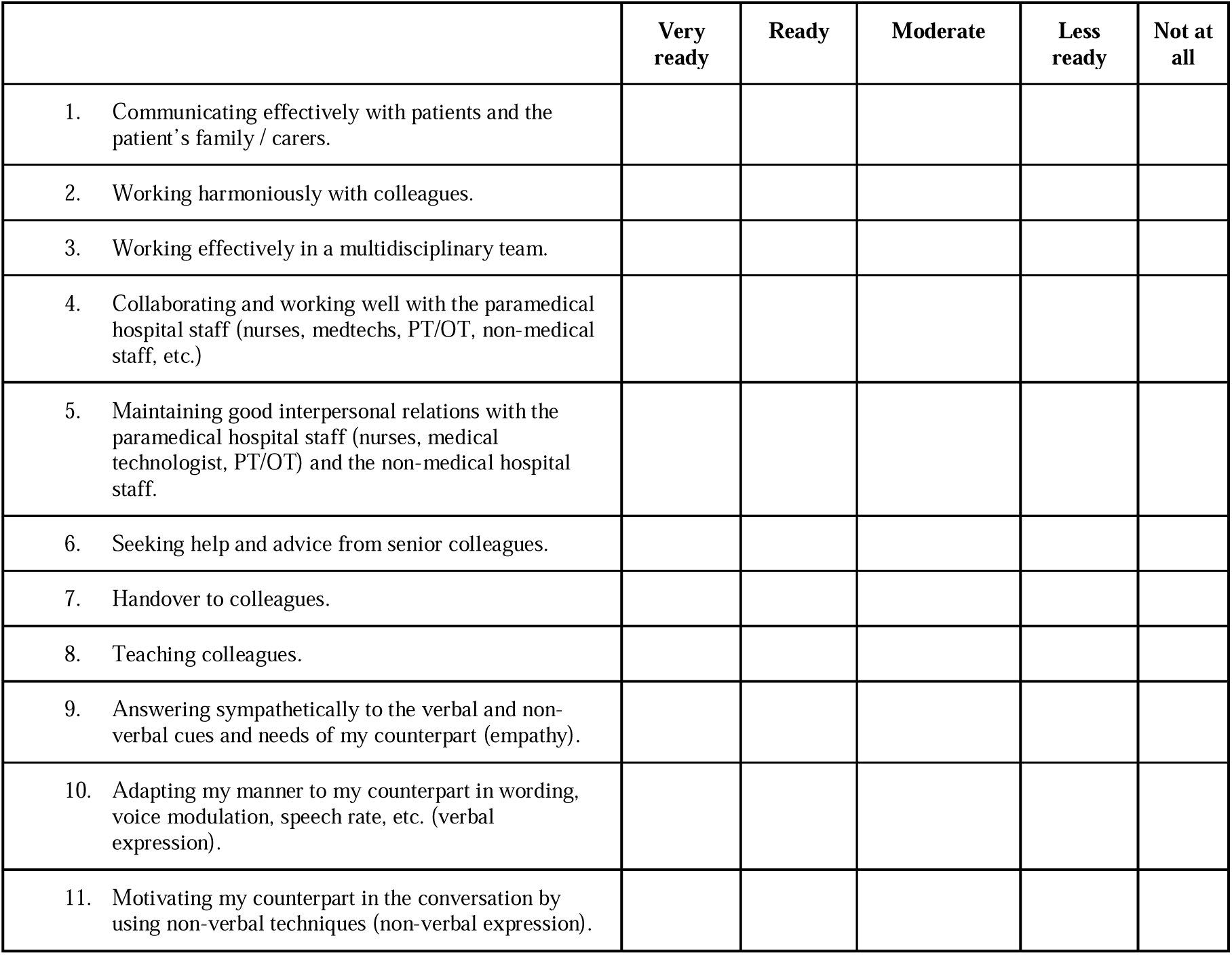

#### D. Clinical Guidelines and Protocols

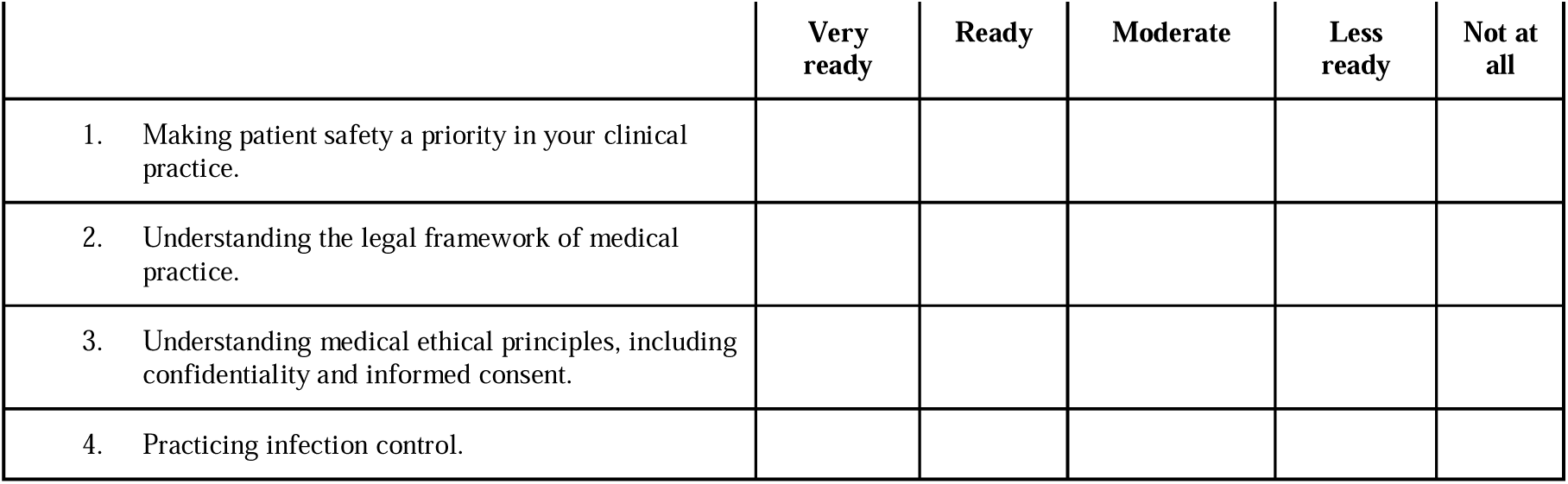

#### E. Personal Development and Wellbeing

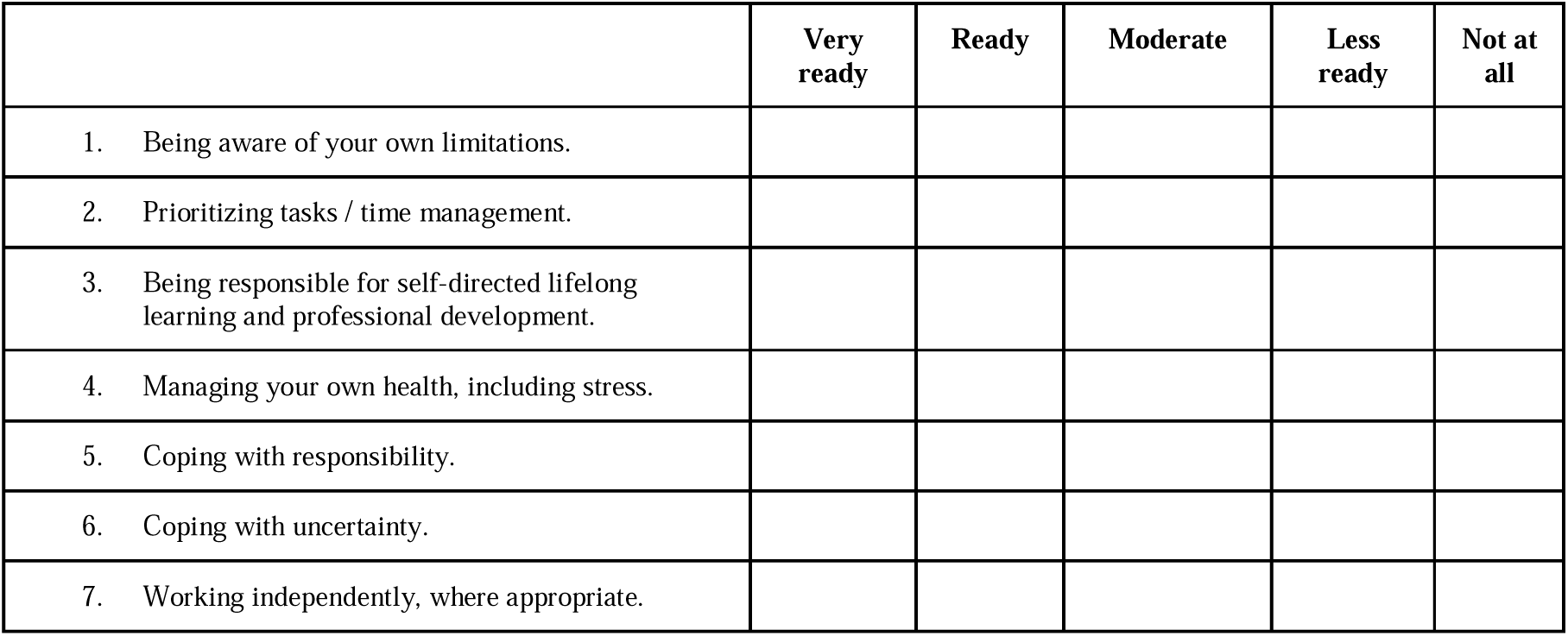

## APPENDIX B. Instrument validity and reliability

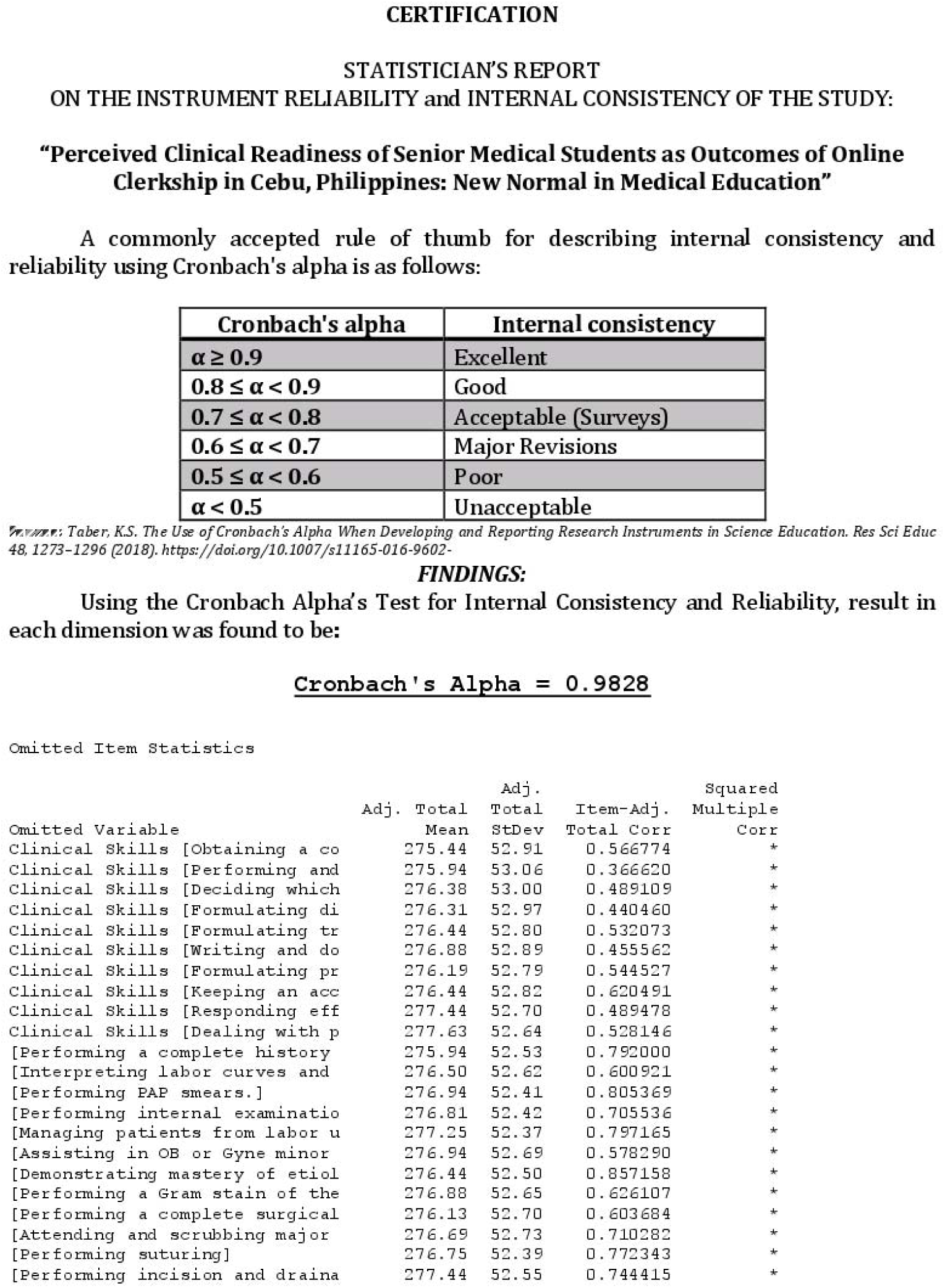
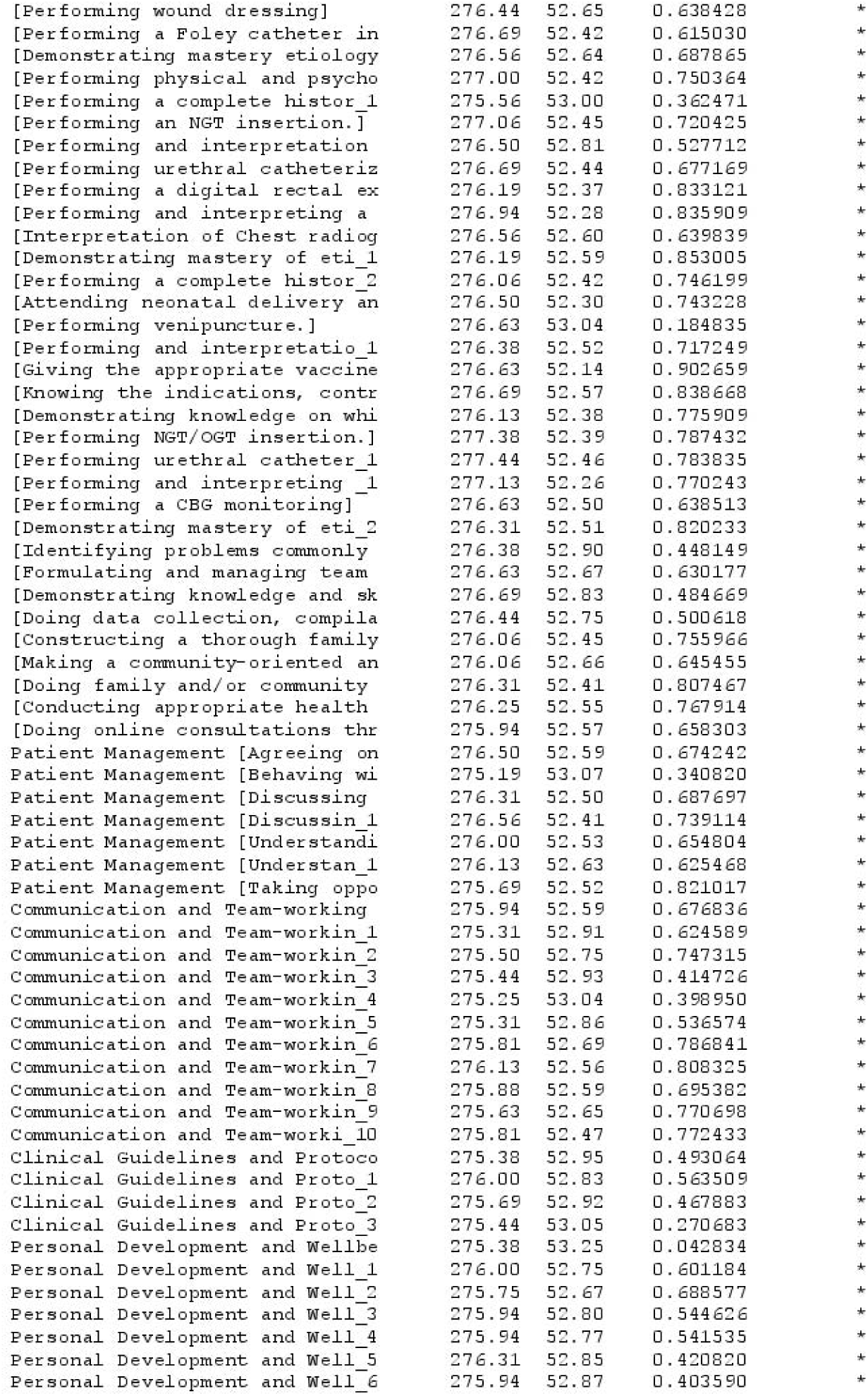
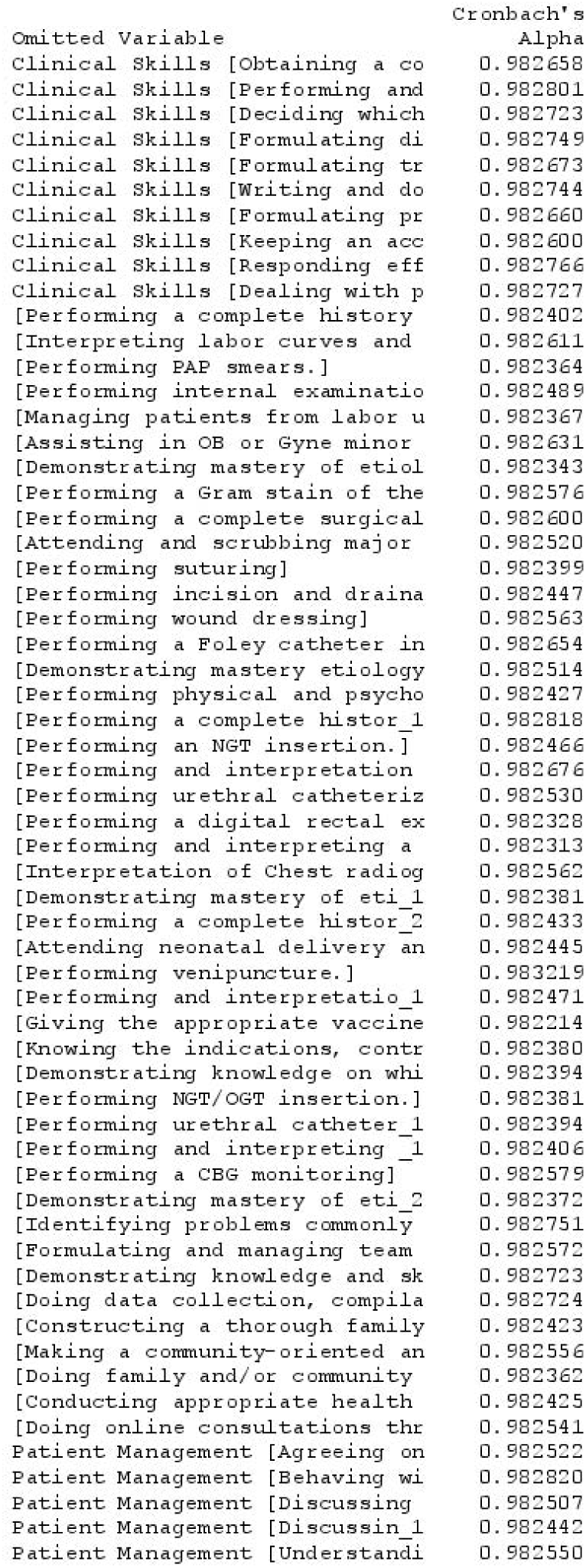
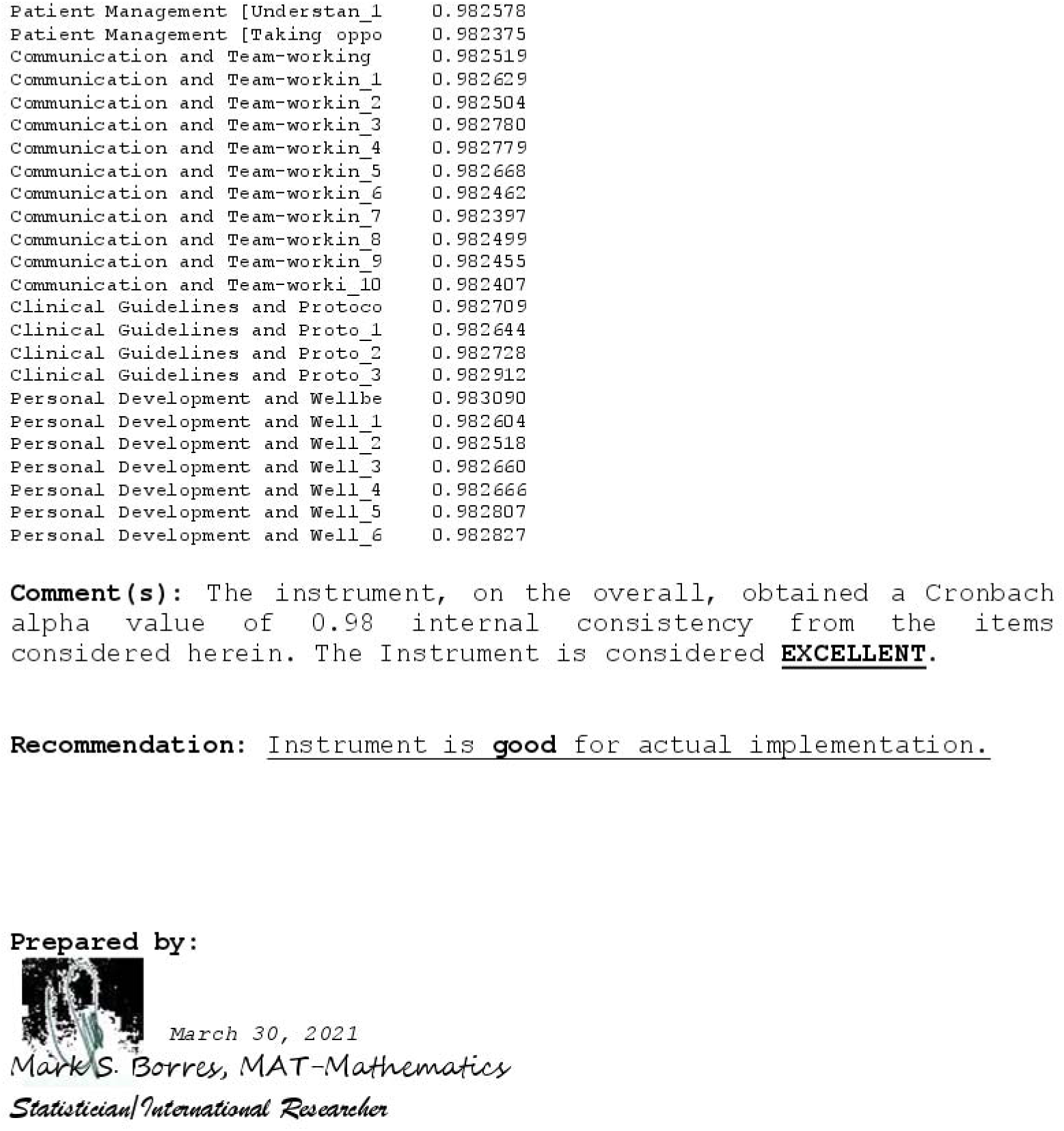

## APPENDIX C. Informed Consent

I understand that I am being invited to participate in a survey/questionnaire activity that forms part of required coursework in Cebu Institute of Medicine. It is my understanding that this survey/questionnaire has been designed to gather information about the Perceived Clinical Readiness of Senior Medical Students. Outcomes of Online Clerkship in the Philippines. The findings of this study will serve as a stepping stone in advancing health care setting in the Philippines that will benefit the following:

### Medical students

This study will allow medical students to evaluate what clinical skills they have acquired so far in the practice of medicine and to assess their level of mastery and confidence when asked to perform the following procedures during return demonstration and in the actual hospital setting. From there, students can have an insight on what areas need improvement and how to overcome the challenges of online learning.

### The community

Conducting this study will also make the community aware that containment of the virus and rapid decline of cases not only aids the healthcare workers, but it also encompasses the rehabilitation of different sectors, most especially, the education department. This will allow continuation of physician-to-patient interaction and honing of fundamental hands-on clinical skills.

I understand that my participation in this project is completely voluntary and that I am free to decline to participate, without consequence, at any time prior to or at any point during the activity. I understand that I may skip any question that I do not wish to answer. I understand that any information I provide will be kept confidential, used only for the purposes of completing this study. Any report of this research that is made available to the public will not include my name or any other individual information by which I could be identified.

All survey/questionnaire responses, notes, and records will be kept in a secure environment. I will also be provided with the raw data or the completed research output at my request.

I also understand that there are no risks and discomforts involved in participating in this activity.

I have read the information above. Completing this survey by answering the questionnaire and signing below indicates that I am 18 years of age or older and indicates my consent to participate in this research.

Signature:_________________________

Date:______________________________

Please keep a copy of this consent form for your records. If you have other questions concerning your participation in this project, please contact us at:

Cell Phone number: +63 917 702 9753

## APPENDIX D. Template for the Letter of Intent to the Schools

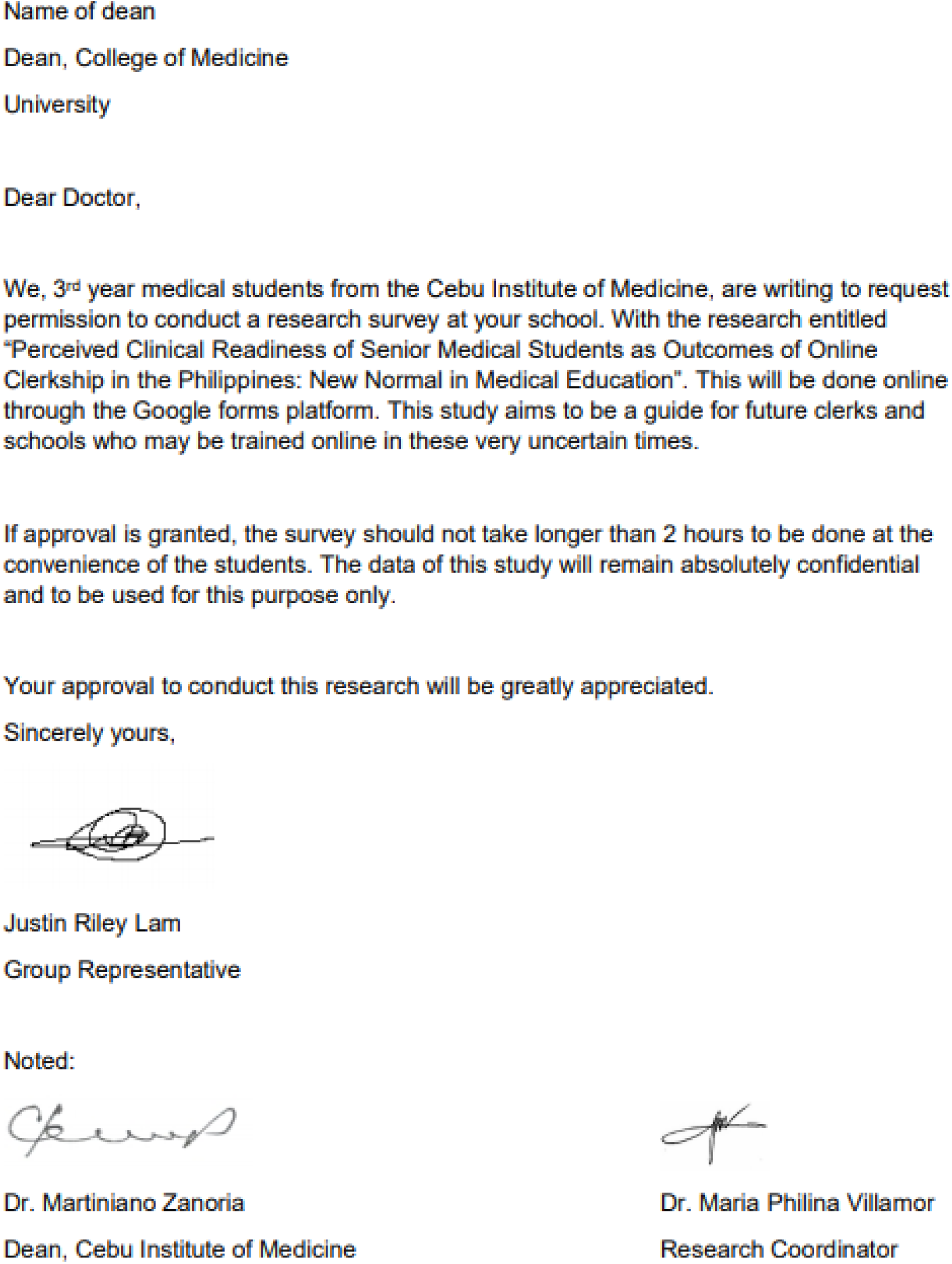

