## Supplementary figures and images for "Perceived Clinical Readiness of Senior Medical Students as Outcomes of Online Clerkship in the Philippines: New Normal in Medical Education"

### IRB

**APPENDIX D**. Letter of Approval from Institutional Review Board (IRB)

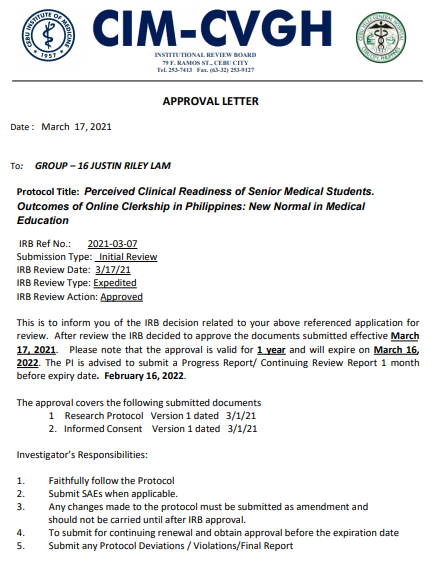

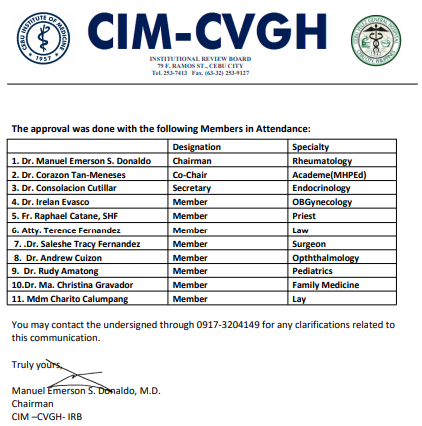
